# Systematic proteomics in Autosomal dominant Alzheimer’s disease reveals decades-early changes of CSF proteins in neuronal death, and immune pathways

**DOI:** 10.1101/2024.01.12.24301242

**Authors:** Yuanyuan Shen, Muhammad Ali, Jigyasha Timsina, Ciyang Wang, Anh Do, Daniel Western, Menghan Liu, Priyanka Gorijala, John Budde, Haiyan Liu, Brian Gordon, Eric McDade, John C. Morris, Jorge J. Llibre-Guerra, Randall J. Bateman, Nelly Joseph-Mathurin, Richard J. Perrin, Dario Maschi, Tony Wyss-Coray, Pau Pastor, Alison Goate, Alan E. Renton, Ezequiel I. Surace, Erik C. B. Johnson, Allan I. Levey, Ignacio Alvarez, Johannes Levin, John M. Ringman, Ricardo Francisco Allegri, Nicholas Seyfried, Gregg S. Day, Qisi Wu, M. Victoria Fernández, Dominantly Inherited Alzheimer Network, Laura Ibanez, Yun Ju Sung, Carlos Cruchaga

**Author notes:** Corresponding author: To whom correspondence should be addressed: Carlos Cruchaga, PhD, Washington University, School of Medicine, 425 S. Euclid Ave. BJC Institute of Heath. Box 8134, St. Louis, MO 63110.

## Abstract

**Background:** To date, there is no high throughput proteomic study in the context of Autosomal Dominant Alzheimer’s disease (ADAD). Here, we aimed to characterize early CSF proteome changes in ADAD and leverage them as potential biomarkers for disease monitoring and therapeutic strategies.

**Methods:** We utilized Somascan® 7K assay to quantify protein levels in the CSF from 291 mutation carriers (MCs) and 185 non-carriers (NCs). We employed a multi-layer regression model to identify proteins with different pseudo-trajectories between MCs and NCs. We replicated the results using publicly available ADAD datasets as well as proteomic data from sporadic Alzheimer’s disease (sAD). To biologically contextualize the results, we performed network and pathway enrichment analyses. Machine learning was applied to create and validate predictive models.

**Findings:** We identified 125 proteins with significantly different pseudo-trajectories between MCs and NCs. Twelve proteins showed changes even before the traditional AD biomarkers (Aβ42, tau, ptau). These 125 proteins belong to three different modules that are associated with age at onset: 1) early stage module associated with stress response, glutamate metabolism, and mitochondria damage; 2) the middle stage module, enriched in neuronal death and apoptosis; and 3) the presymptomatic stage module was characterized by changes in microglia, and cell-to-cell communication processes, indicating an attempt of rebuilding and establishing new connections to maintain functionality. Machine learning identified a subset of nine proteins that can differentiate MCs from NCs better than traditional AD biomarkers (AUC>0.89).

**Interpretation:** Our findings comprehensively described early proteomic changes associated with ADAD and captured specific biological processes that happen in the early phases of the disease, fifteen to five years before clinical onset. We identified a small subset of proteins with the potentials to become therapy-monitoring biomarkers of ADAD MCs.

**Funding:** Proteomic data generation was supported by NIH: RF1AG044546

## Introduction

Autosomal dominant Alzheimer’s disease (ADAD) accounts for approximately 1% of all AD cases.^1^ ADAD is characterized by the presence of autosomal dominant mutations in amyloid precursor protein (*APP*), presenilin-1 (*PSEN1*), or presenilin-2 (*PSEN2*).^1,2^ This rare form of AD has been instrumental in elucidating critical pathological mechanisms that underlie the disease and the temporal progression of brain changes associated with AD.^3^ Thus, a comprehensive study of the cerebrospinal fluid (CSF) proteomic changes in this rare form of the disease can help advance our understanding of its intricate pathophysiology and contribute to the identification of novel biomarkers and potential therapeutic strategies.

The Dominantly Inherited Alzheimer Network (DIAN) Observational Study is a worldwide effort to study individuals with ADAD mutations, aiming to understand the natural progression of ADAD. It involves longitudinal assessments including imaging, cognitive evaluations, and fluid collection including CSF and plasma.^4^ Traditional CSF AD biomarkers, such as amyloid β42 (Aβ42), total Tau (Tau), and phosphorylated-tau181 (pTau) are measured regularly and have demonstrated their analytical validity in ADAD as well as sporadic Alzheimer’s disease (sAD).^5^ Additionally, several studies support the correlation between CSF biomarker levels and neuropathological changes associated with AD onset and progression.^5–7^ In fact, CSF biomarkers change earlier in the disease course than amyloid or tau in positron emission tomography (PET) imaging.^8^ Moreover, CSF biomarkers provide more cost-effective, faster, and simpler results in clinical settings than those based on imaging techniques.^7^ Although proteome-based biomarkers in the plasma for ADAD and sAD are being identified, CSF, which is in direct contact with the brain, is relatively unaffected by proteins from other organs.^9^ In ADAD, mutation carriers (MCs) from the same family tend to exhibit similar age at symptom onset (AAO), and therefore, it is possible to calculate the “estimated year of onset (EYO)” for each family member by extrapolating from the known AAO in individuals who share the same mutation.^10^ EYOs are determined by subtracting the age at study assessment minus the mean mutation AAO associated with their specific mutation. ^10^

ADAD exhibits several similarities to sAD, including phenotype, clinical progression, and neuropathology.^1,11,12^ However, it is essential to note that the etiology and onset mechanisms of these two AD forms follow distinct patterns.^13^ The first published unbiased proteomic study of CSF in 14 carriers of ADAD mutations identified 600 proteins, 56 of which differed between MCs and NCs. ^14^ Of these, 40 proteins displayed alterations during the presymptomatic stage of the disease and fourteen of these proteins had been previously described to be altered in sAD. In a recent proteomic study that included 22 MCs and 20 NCs individuals, 66 CSF proteins associated with ADAD were identified.^15^ When comparing with sAD (n=531), they demonstrated that ADAD indeed presented biomolecular similarities with sAD, and in fact, the common proteins were dysregulated in both ADAD and sAD. However, the predictive value of those proteins was not explored. In another recent proteomic study, researchers analyzed 59 CSF proteins in 286 MCs and 184 NCs samples.^16^ However, these 59 proteins were pre-selected based on previous brain studies. Out of 59, 33 proteins were associated significantly with ADAD status after correcting by EYO.

In recent years, there has been a significant increase in the volume of AD proteomic research. These studies have explored protein variances in AD across various brain regions^17–19^ and tissue fractions,^20–24^ such as those enriched in insoluble materials, synaptic components, membranes, or blood vessels. Furthermore, specialized proteomic analyses have been conducted on a small scale but has been able to capture some actual biological substrates and goes beyond the traditional neuropathological features like amyloid plaques, neurofibrillary tangle, and cerebral amyloid angiopathy, ^25–31^ leading to a better biological understanding of the disease as well as revealing potential new therapeutic targets. Each study has individually contributed valuable insights into the pathogenesis, revealing novel potential drug targets and biomarkers but mostly in sAD. However, when evaluating individual studies, including the previous published ADAD studies, they shared some common limitations such as limited sample size and a small number of investigated proteins. Moreover, it is worth noting that previous studies were focused on identifying proteins associated with mutation status, not disease onset. Although some were corrected by the EYO,^16^ EYO information has neither been fully leveraged to identify proteins that show different trajectories between MCs and NCs, nor to determine the time in which protein changes occur in relation to the clinical onset. Overall, these limitations highlight the need for an unbiased large-scale and high-throughput study to identify early proteomic changes in the ADAD.

Therefore, in this study, we utilized a deep and unbiased biofluids proteomic profiling (covering CSF and plasma) and measured 6,163 proteins in 476 CSF samples (291 MCs and 185 NCs) and 6,022 proteins in 538 plasma samples (325 MCs and 213 NCs) from DIAN by Somascan® 7K Platform. High-throughput unbiased proteomics goes beyond standard AD biomarkers studies or neuroimaging as proteomics is getting at the actual biological substrates, which can lead to a much better biological understanding of the disease as well as revealing potential new therapeutic targets.

To conduct this study, we used a novel approach that leverages the estimated age at onset to assess pseudo-trajectories (using cross-sectional data to simulate longitudinal data) and identified proteins with significantly different coefficient-estimates between MCs and NCs. We subsequently determined the exact time when those pseudo-trajectories become divergent in relation to the EYO. We applied network and pathway-enrichment analysis to create clusters of proteins that share functional relationships, leading to the identification of novel mechanisms implicated in disease. Finally, we leveraged those dysregulated proteins to create Aβ- and pTau-independent predictive models. Our overarching goal was to pinpoint potential biomarkers that could aid in monitoring disease progression, assessing treatment effectiveness, and designing well-informed therapeutic strategies.

## Methods

### ADAD cohort

DIAN is a long-term observational study that employs standardized clinical and cognitive assessments (Clinical Dementia Rating®(CDR)®), neuropsychological testing, imaging modalities (magnetic resonance imaging (MRI), PIB-PET, and 18F-FDG),^32^ and collects biological fluids (blood and CSF). The main goal of this cohort is to detect alterations in individuals who possess known gene mutations that cause AD, including pre-symptomatic and symptomatic mutation carriers. We applied the Somascan® 7K panel to analyze the CSF proteome from 291 MCs, 185 NCs, and the plasma proteome from 325 MCs and 213 NCs from DIAN participants. In total, there were 448 participants that have both CSF and plasma proteomics data.

CSF samples were distributed as follows: 291 MCs [216 (74%) *PSEN1*, 23 (8%) *PSEN2,* and 52 (18%) *APP*]. Of these, 105 (36%) were symptomatic MCs, and 186 (64%) were presymptomatic MCs. The 108 NCs in the study were recruited from family members associated with MCs. The mean age of MCs was 40.0 years; 55% were females, with a mean EYO of −6.6 ±10.4 years (yrs), and 29% of MCs carried at least one APOE ε4 allele. The mean age of NCs was 39.6 years, 59% were females, the mean EYO was −7.8 ± 12.5 yrs, and 33% of NCs presented at least one APOE ε4 allele. Age, sex, EYO and APOE ε4 allele did not show any significant difference between MCs and NCs in CSF (**Table 1**).

**Table 1.**
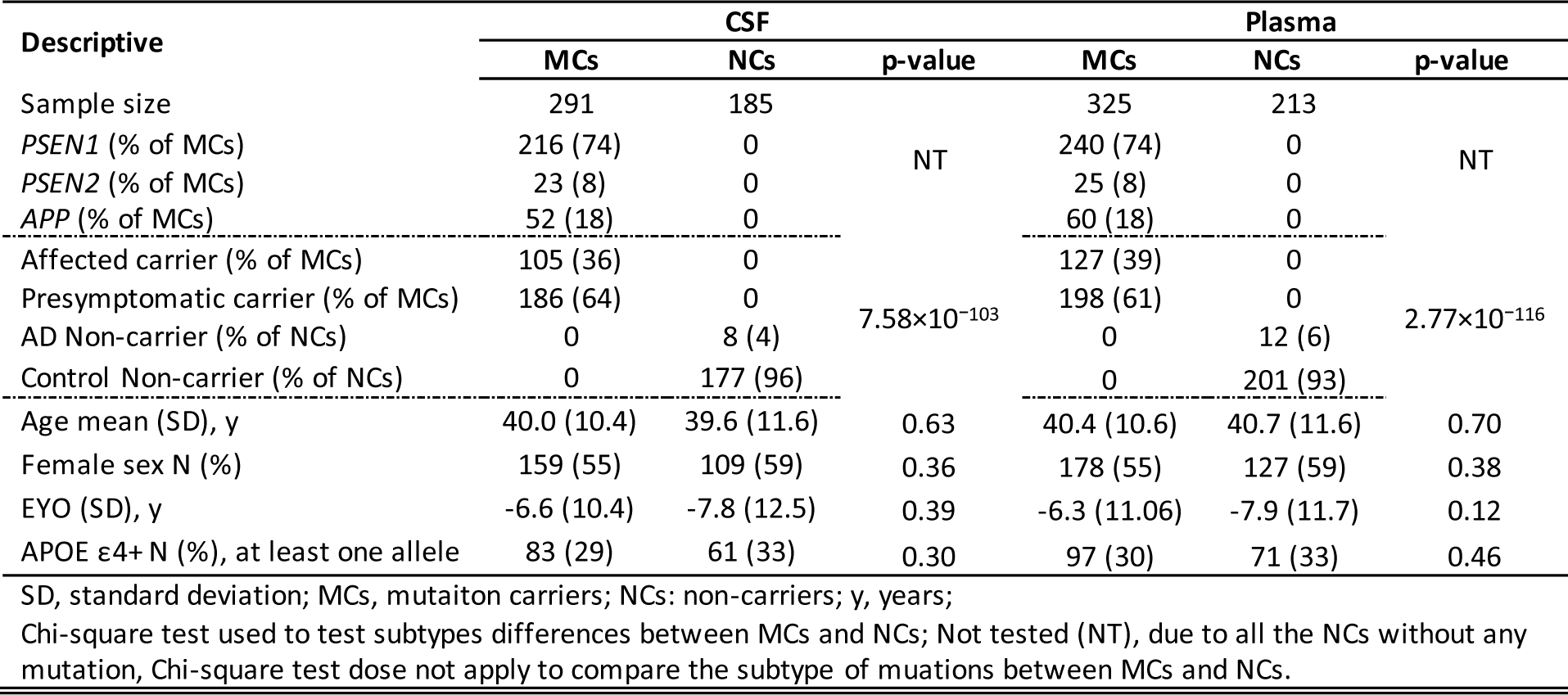
Demographics of participants of CSF and plasma in DIAN.

| Descriptive | CSF |  |  | Plasma |  |  |
| --- | --- | --- | --- | --- | --- | --- |
|  | MCs | NCs | p-value | MCs | NCs | p-value |
| Sample size | 291 | 185 |  | 325 | 213 |  |
| *PSEN1* (% of MCs) | 216 (74) | 0 | NT | 240 (74) | 0 | NT |
| *PSEN2* (% of MCs) | 23 (8) | 0 |  | 25 (8) | 0 |  |
| *APP* (% of MCs) | 52 (18) | 0 |  | 60 (18) | 0 |  |
| Affected carrier (% of MCs) | 105 (36) | 0 |  | 127 (39) | 0 |  |
| Presymptomatic carrier (% of MCs) | 186 (64) | 0 | $7.58 \times 10^{-103}$ | 198 (61) | 0 | $2.77 \times 10^{-116}$ |
| AD Non-carrier (% of NCs) | 0 | 8 (4) |  | 0 | 12 (6) |  |
| Control Non-carrier (% of NCs) | 0 | 177 (96) |  | 0 | 201 (93) |  |
| Age mean (SD), y | 40.0 (10.4) | 39.6 (11.6) | 0.63 | 40.4 (10.6) | 40.7 (11.6) | 0.70 |
| Female sex N (%) | 159 (55) | 109 (59) | 0.36 | 178 (55) | 127 (59) | 0.38 |
| EYO (SD), y | -6.6 (10.4) | -7.8 (12.5) | 0.39 | -6.3 (11.06) | -7.9 (11.7) | 0.12 |
| APOE $\epsilon 4$+ N (%), at least one allele | 83 (29) | 61 (33) | 0.30 | 97 (30) | 71 (33) | 0.46 |

SD, standard deviation; MCs, mutation carriers; NCs: non-carriers; y, years;
Chi-square test used to test subtypes differences between MCs and NCs; Not tested (NT), due to all the NCs without any mutation, Chi-square test does not apply to compare the subtype of mutations between MCs and NCs.

Plasma was available for 325 MCs [240 (74%) *PSEN1*, 25 (8%) *PSEN2,* and 60 (18%) *APP*]. Of those, 129 (39%) were asymptomatic MCs, and 196 (61%) were presymptomatic MCs. The mean age of MCs was 40.4 years; with a mean EYO of −6.3 ±11.06 yrs, 30% carried at least one APOE ε4 allele, and 55% were females. There were 213 NCs with a mean age of 40.7 years; 59% were females, with the mean of −7.9 ± 11.7 yrs, and 33% of NCs carried at least one APOE ε4 allele. There was no significant difference observed in the distribution of sex, age at draw, EYO, and frequency of APOE ε4 carriers as well in the individual with plasma samples and at the specific blood draw used in this study.

The Institutional Review Board of Washington University School of Medicine in St. Louis approved the study, and research was performed following the approved protocols. All human studies were conducted under the supervisory review and approval of the Washington University in St. Louis institutional review board. Participants or their caregivers provided informed consent by their respective local institutional review boards.

### Clinical assessment and EYO in ADAD

Dementia was evaluated using the CDR®, with clinical assessors blinded to participants’ mutation status. The EYO for a participant was computed during each assessment, factoring in their age at the visit and the anticipated period of dementia symptom onset specific to their mutation. ^10^ This anticipated onset age for a given mutation was established by averaging reported symptom onset ages among individuals sharing the same mutation. When the mutation-specific expected age of symptom onset was unavailable, the EYO was derived from the age of cognitive decline onset in the participant’s parents or family members, as determined through a thorough semi-structured interview using comprehensive historical data. It is crucial to note that the EYO calculation procedure remained consistent for both MCs and NCs. In the present study, we leveraged the DIAN-EYO, which enhances EYO accuracy by integrating an individual’s actual decline age into the EYO determination process rather than solely relying on the mean mutation or parental/familial age at onset. ^10^ For simplification, EYO stands for DIAN-EYO.^10^ Mutation status was determined using a PCR-based amplification of the relevant exon, followed by Sanger sequencing.^2^

### Sporadic AD cohorts

To characterize ADAD proteomics in comparison to sAD, we combined four different sAD cohorts (Charles F. and Joanne Knight Alzheimer Disease Research Center (Knight-ADRC), Alzheimer’s Disease Neuroimaging Initiative (ADNI), Fundació ACE Alzheimer Center Barcelona, and Barcelona-1; see supplementary materials) with available CSF proteomic data, reaching a total of 1,763 samples. We grouped the sAD cohorts based on the ATN classification (where A^+^T^+^ and A^−^T^−^ serve as proxies for sAD and controls).^33^ However, the lack of consensus in universal biomarker cutoffs is a major caveat as biomarker levels, and subsequently their cutoffs for dichotomization, can be influenced by the technique of measurement. Thus, we utilized Gaussian mixture models to dichotomize quantitative Aβ42 and pTau measures into positive and negative groups and applied separately for each sAD cohort. ^34,35^ Individuals with low CSF Aβ42 and high pTau levels were classified as amyloid/tau positive (A^+^T^+^). Individuals with high Aβ42 and low pTau levels were defined as controls (A^−^T^−^). Detail description of the dichotomization and cut-off determination for each cohort can be found elsewhere ^36^ and in the supplementary materials.

### CSF proteomic data collection, processing, and quality control (QC)

The Somascan® 7K Platform is the aptamer-based technology with modified DNA aptamers (SOMAmers), that measures relative protein levels in a multiplex fashion. ^37^ This technology has several advantages compared to other classic approaches (ELISA and MS) because it theoretically suffers less from the challenges of dynamic range and missing value. CSF samples were collected from each cohort via lumbar puncture in the morning following an overnight fasting. All samples underwent the same preparation and processing protocols and were stored at −80 °C. We started to measure a total of 7,584 aptamers across 495 DIAN samples and 1,763 sAD and controls using SomaLogic’s Somascan® Platform. The samples were sent together to SomaLogic to minimize batch effects and randomly distributed across plates. Protein abundance levels were measured using the SomaLogic aptamer-based Somascan® platform, which utilizes a multiplexed-based single-stranded DNA aptamer assay for protein quantification. The protein levels were reported as relative intensity units (RFU or Relative Fluorescence Unit). For data normalization, SomaLogic conducted an initial step using hybridization controls to account for intra-plate variability and median signal to address inter-plate variability.^38^ Additionally, SomaLogic performed an additional normalization step by comparing the data against an external reference to control for biological variation.^39^ To ensure data quality, individual-level quality control (QC) was carried out to detect and exclude outlier aptamers and samples, as described previously. ^34,39^

In summary, after rigorous quality control, 476 samples from the DIAN study and 1,763 samples from sporadic Alzheimer’s disease (sAD) were retained for further analyses. These samples were examined using 7,029 protein aptamers in sAD, among which 23 protein aptamers did not have corresponding protein. In the DIAN cohort, 7,008 aptamers were examined, and two protein aptamers targeted HIV proteins (HIV-2 Rev, C34 gp41 HIV Fragment). These two HIV proteins were not analyzed in sAD. Overall, in DIAN and sAD cohorts, there were 845 and 866 aptamers targeting more than one protein, respectively; and in total, we included 6,163 unique proteins in the analysis.

In addition, levels of CSF Aβ42, total Tau (Tau), and pTau, not present on the SomaLogic panel were also available for the DIAN and sAD samples that were measured using Lumipulse® G Assays (Fujirebio) on the Lumipulse® G platform.^40,41^

### DIAN plasma proteomics data collection, processing, and quality control

Blood samples were collected at the time of the visit, immediately centrifuged, and the plasma stored at − 80°C. Clinical status (case-control) was determined by the CDR® at the time of draw. We measured 7,584 aptamers in 580 plasma samples from DIAN participants using SomaLogic’s Somascan® Platform. Initial data normalization and QC was performed as described above. Ultimately, 564 samples and 6,985 aptamers targeting 6,022 unique proteins were kept for downstream analysis. In the subsequent description, we will only talk about unique proteins for both CSF and plasma, instead of protein aptamers.

### Statistical analysis

#### Cohorts Demographics

We processed demographic characteristics using GraphPad Prism 9. To compare cohorts characteristics between MCs’ and NCs’ groups we used unpaired student t-tests and Chi-squared tests for continuous and binary variables, respectively. Continuous variables were reported as mean ± standard deviation. P values of <0.05 after the false discovery rate correction method was considered significant.

#### Differential pseudo-trajectory analysis

To study how alterations in protein levels changes over time, we employed a multi-layer regression model to infer the pseudo-temporal trajectory from the cross-sectional proteomic data, called pseudo-trajectory analysis. This model constructed a more comprehensive framework, capable of capturing proteins with significant pseudo-trajectory differences between MCs and NCs. In the pseudo-trajectory calculation model, we incorporated the first two surrogate variables (SVs) to correct for unmeasured heterogeneity. These SVs were generated by applying the *sva()* function within the R (version 4.1.3) package *sva* (version 3.42.0). Pseudo-trajectory calculations were performed in three steps: (1) we used a linear regression, *lm()* function from the *stats* package, to build the model with log 10-transformed proteins as the dependent variable, EYO as the independent variable, sex, and the first two SV as covariates for MCs and NCs, separately (Formula 1; supplementary materials); (2) we compared the coefficient-estimates (β) of EYOs between MCs and NCs (Formula 2); and (3) we calculated when the pseudo-trajectories significantly deviate from each other in EYO by student t-test (Formula 3, 4). All the formulas are presented in the “Differential pseudo-trajectory analysis” section in the supplementary materials. Bonferroni correction was used for the CSF analyses and False Discovery Rate (FDR) correction (FDR p < 0.05) for plasma, as the plasma analyses were exploratory.

#### Pseudo-trajectory intersections in ADAD

For each protein that displayed substantial differences, we calculated the pseudo-trajectory intersection point between MCs and NCs. This was done to calculate the exact moment when the protein levels between these two groups started to diverge significantly. We followed a two-step process to derive pseudo-trajectory intersections for each protein (Model 2, formula 3 and 4; supplementary materials). In the initial step, we utilized the *predict()* function to generate the predicted 95% confidence intervals (CI) for MCs and NCs independently (Formula 5). In the second step, we extracted the lower bound and upper bound values from the predicted 95% CIs of both MCs and NCs, assembling them into a new matrix. We established another linear regression model from this new matrix by selecting either the predicted lower bound or upper bound values for significant protein based on each protein’s specific β. If the β for a particular protein were > 0, indicating that its protein level was higher in MCs than NCs, we opted for the lower bound from the MCs matrix and the upper bound from the NCs matrix. Conversely, if the β < 0, indicating that the protein level was higher in NCs than MCs, we selected the upper bound from the MCs matrix and the lower bound from the NCs matrix. Following this selection, we extracted the two βs from the new linear regression models (Formula 6).

#### ADAD protein change and sAD protein change analysis in ADAD and sAD

Protein level changes between mutation status groups (MCs vs. NCs) in the DIAN cohort and between ATN status in sporadic late-onset AD cohorts (A^+^T^+^ vs. A^−^T^−^) were identified using the linear regression model. In these models, the log 10-transformed protein levels served as the dependent variable, mutation or ATN and the group status as the independent variable, and age at CSF draw, sex and the first two SVs included as covariates (see supplementary materials).

#### Proteomic comparison across studies

To compare the analyses and results across cohorts and analyses, we evaluated the consistency of CSF ADAD results with other studies by comparing the effect sizes for each protein. These comparisons included (1) another two publicly available CSF ADAD studies,^15,16^ (2) the analyses for CSF in sporadic AD and (3) DIAN plasma ADAD results. We obtained correlation (*R2*) and significance (p value) by performing correlation tests using Pearson’s correlation test from the *cor.test()* function in R to test effect sizes of studies. The prediction line and regression line’s 95% confidence interval were utilized to assess the effect size of significant proteins between ADAD and sAD.

#### Predictive models

We used the least absolute shrinkage and selection operator (LASSO – L1 regularization) regression model with five-fold cross-validation to identify the minimum set of the most informative proteins to build a predictive model for ADAD mutations status. We used the *train()* function in the *caret* R package version 6.0 to employ the LASSO regression model. Starting from an initial set of 125 differential pseudo-trajectory proteins, we identified a final subset of nine to create the ADAD-specific predictive model. We used the traditional AD markers for benchmarking the newly generated model.

The predictive model was trained using 70% of complete dataset (discovery data) by setting random seeds to split DIAN cohort, and tested on the remaining 30% of the data (replication data). The sensitivity (true-positive rate) and specificity (true-negative rate) of the developed ADAD predictive model were assessed by plotting the receiver operator characteristic (ROC) curves using *pROC* R package version 1.18.2.^42^ To further evaluate the performance of these proteins, we generated areas under the curves (AUC) and estimated the positive predictive value (PPV) and negative predictive value (NPV) based on Youden’s J statistic optimal cut-off using “cords” function in the pROC R package.^43^

### Functional Analysis

#### Pathway enrichment analysis

We performed pathway enrichment analysis to identify biological functions of proteins using *ClusterProfiler* R package version 4.8.0.^44^ Default parameters were used for the analysis, with FDR P-value <0.05 to define the significantly enriched pathways; a minimum count of three were collected and grouped into clusters based on their membership similarities. In each module, all the enriched pathways were grouped into different categories. This categorization was based on the “Event Hierarchy” within the Reactome database, a decision driven by the recognition of numerous repetitive branch pathways that were enriched with the same proteins. We used *enrichDGN()* function to examine the associations of identified proteins with other diseases based on the DisGeNET database.^45^ Biological pathway enrichment analysis was performed using the *enrichPathway()* function within the *ClusterProfiler* environment and the Reactome database.^46^ The top 20 pathways were shown graphically using the *barplot() and emapplot()* functions from the *ClusterProfiler* package. The STRING database was used to build the protein-protein interactions and describe the subcellular structures and macromolecular complexes by integrating with the cellular component ontology database.^47^ FDR P-value <0.05 was used to define the significant enriched protein-protein interaction. Druggable genome was used to map the targetable genes in modules.^48^

#### Weighted Gene Correlation Network Analysis (WGCNA)

We used the WGCNA R package (Version 1.72.1)^49^ algorithm to generate a central network of co-expression modules for identified proteins. The *pickSoftThreshold()* function was leveraged to establish the soft threshold power at 12, with a minimum module size of five. The resulting correlation matrix was transformed into a signed adjacency matrix, which calculates a topological overlap matrix (TOM), representing expression similarity across samples for all proteins in the network. This approach uses hierarchical clustering analysis as one minus TOM, and dynamic tree cutting leads to module identification. Following construction, module eigenprotein (ME) values were defined —representative abundance values for a module explaining modular protein covariance. Pearson correlation between proteins and MEs was used as a module membership measure, defined as kME.

#### Cell type Enrichment Analysis

The cell marker data used in our research was sourced from the study conducted by Zhang Y, et al. ^50^ This data included markers specific to human astrocytes, neurons, oligodendrocytes, microglia/macrophages, and endothelial cells. The purpose of using this data was to evaluate the specificity of the proteins included in the Somascan® 7K panel in relation to these relevant cell types. The data downloaded contained multiple subtypes of astrocytes; we focused only on human mature astrocytes for our analysis. For each cell type, we determined the average expression level across individuals for each gene. We then added the averages from each cell type to get a total expression level for that gene across the five cell types. We then calculated the percentage of the total expression that each cell type contributed. A gene was reported to be cell-type specific if the percentage of its full expression contributed by the top cell type was 1.5× higher than the second top cell type. To determine enrichment, we first matched each of the proteins in the Somascan® 7K platform to their Gene Symbols using the SomaLogic-provided documentation. Using the above ratio-based strategy, we determined the cell-type specificity for all Gene Symbols and counted the number of genes specific to each cell type. In total, 5,743 proteins had host genes included in the cell-type expression data. For each protein subset, we determined the number of related genes specific to each cell type. We then tested for enrichment by comparing the background gene with fold count change. Log2 transformed fold count change was used to plot the cell type enrichment.

#### Data visualization

Data visualization plots were mainly generated by ggplot2 R package version 3.4.2. Specifically, the results of the differential proteins analysis in the form of significantly up- and down-regulated proteins were visualized as a volcano plot using the *geom_point()* function. Pseudo-trajectory curve and trajectory intersections were visualized by stat_smooth() function with ‘loess’ method.

## Results

### Study Design

The primary goal of this study was to identify proteins that present early changes in ADAD mutation carriers (MCs) compared to non-carriers (NCs) in both CSF and plasma. We aimed to identify the earliest biomarkers of ADAD, with the potential to provide valuable insights into the pre-symptomatic phases of ADAD and potentially enable early intervention strategies. By leveraging these proteins, our ultimate goal was to contribute to the development of more effective treatment monitoring techniques and propose novel therapeutic strategies for ADAD patients. This research has the capacity to advance our understanding of the disease and improve the prospects for those at risk.

We generated and leveraged large-scale unbiased proteomics from CSF (6,163 proteins) and plasma (6,022 proteins; **Fig. 1**). Our analytical approach centered on utilizing well-designed informatics models, enabling us to extract proteins exhibiting statistically significant differences in their trajectories between MCs and NCs. This method was chosen for its capacity to closely approximate the pseudo-trajectory of protein alterations in these two distinct groups. Subsequently, we pinpointed when these significant protein changes commenced, providing invaluable insights into the disease progression. We applied co-expression network analysis to cluster the significant proteins for a more comprehensive perspective. Further, we conducted a thorough pathway enrichment analysis with multiple resources to enhance our understanding of the biological implications of these significant proteins within their distinct clusters. For the DIAN plasma samples, we utilized the same analytical pipeline that we applied to the CSF. Our goal was to determine whether plasma samples could provide information consistent with that obtained from CSF samples. Crucially, both the CSF and plasma samples were sourced from the same cohort. We then employed publicly accessible ADAD datasets to validate our findings from CSF proteomics. Additionally, we investigated whether the proteins associated with ADAD were also linked to sporadic late-onset Alzheimer’s disease (sAD). In the final phase, we applied the power of the LASSO model on the most representative significant proteins, which could serve as promising biomarkers for ADAD. This project holds substantial promise for advancing our understanding of ADAD and improving diagnostic and therapeutic strategies.

**Figure 1.**
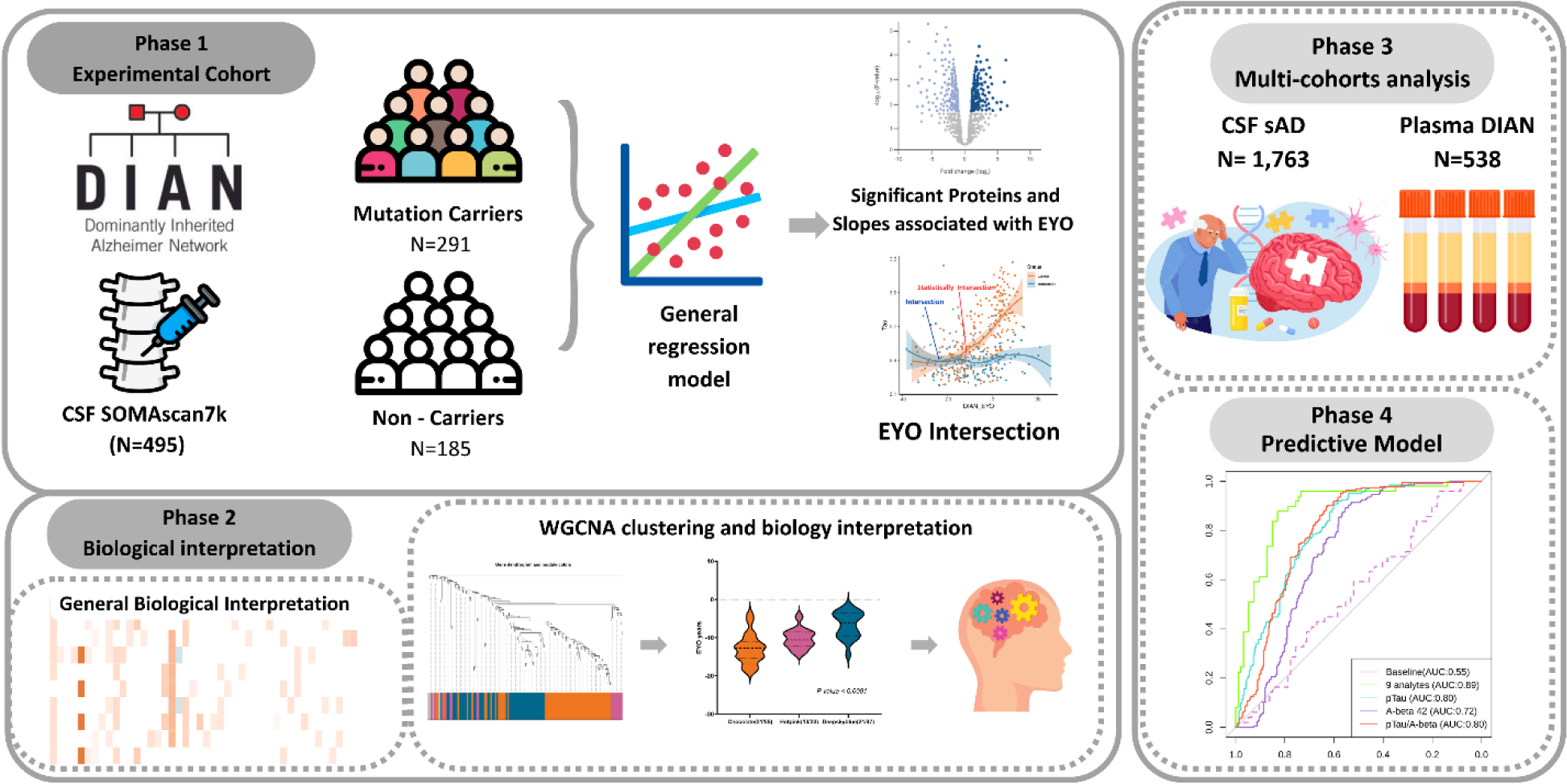
Study overview. In phase 1, proteins measured in the CSF sample were obtained with Somascan®, targeting 6,163 proteins from DIAN. In this stage, the experimental cohort contains 291 MCs and 185 NCs. Differential pseudo - trajectory analyses were performed between MCs and NCs. Trajectory intersections were calculated for significant pseudo-trajectory proteins. Biological functions were identified by protein co-expression network analysis and pathway enrichment. A total of 1,763 sAD CSF samples and 538 DIAN plasma samples wer e analyzed to validate the approach and contextualize the findings. Several publicly available external proteomic datasets were used to validate our findings as well (details in supplementary materials). Last, the LASSO model was used to select significant trajectory proteins and create predictive models for ADAD.

Baseline characteristics of the DIAN participants were summarized in **Table 1**. MCs include symptomatic and presymptomatic DIAN participants with pathogenic variants in one of three ADAD genes *(APP, PSEN1, and PSEN2)*.^51^ Presymptomatic carriers are MCs with CDR® = 0 at CSF draw date. The matched NCs were sourced from the families of the MCs. This control group shares similar genetics background and environmental influences, further enhancing the power of our study to detect reliable proteomic alterations.

### Significantly different pseudo-trajectory protein identification in CSF samples

We analyzed 6,163 proteins that passed quality check from 291 MCs and 185 NCs. Following stringent Bonferroni threshold (p < 7.13×10^−6^), we identified 125 proteins (132 protein aptamers) that exhibited significantly distinct trajectories between MC vs NCs. In addition, 1,069 proteins showed nominal (P < 0.05) significance (**Fig. 2A, Table S1)**. Additionally, Aβ, p-tau, and tau were included for comparison as they are known and well validated AD biomarkers.^40,41^

**Figure 2.**
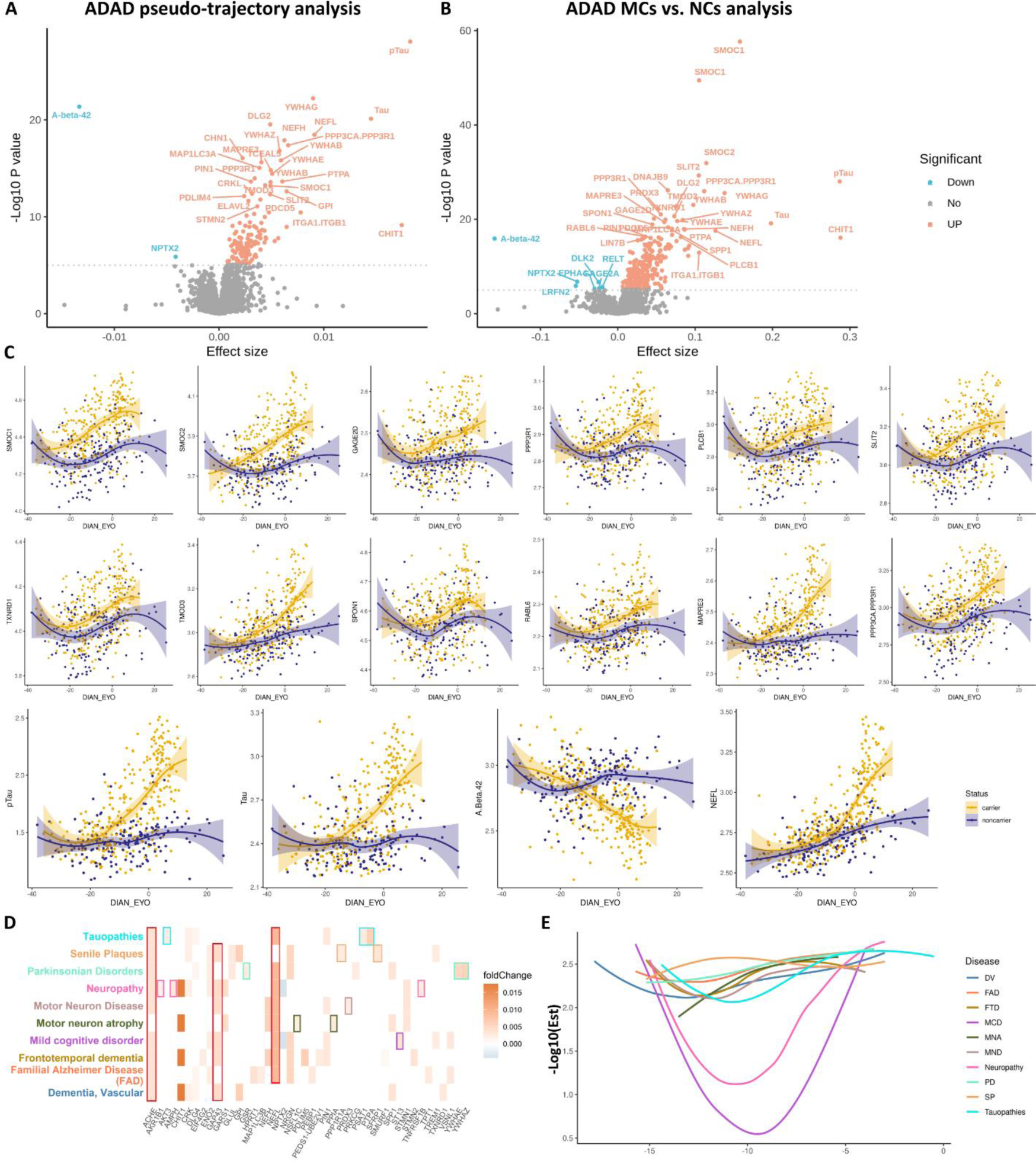
Significant pseudo-trajectory proteins and significant proteins associated with ADAD mutation status in CSF. **(A-B)** Volcano plots displaying the estimate change (x axis) against −log10 statistical differences (y axis) for all tested proteins. The red dots show the significantly upregulated proteins and the blue dots show the significantly downregulated proteins at Bonferroni threshold (p < 7.13×10^−6^).**(A)** Volcano plot for the pseudo-trajectory analyses comparing trajectories between MC and NC; **(B)** Volcano plot for the ADAD mutation status only; **(C)** Twelve significant pseudo-trajectory proteins that changed earlier than Tau, pTau, Aβ42. The ‘loess’ parameter in the plotting was applied to reflect the accuracy of the protein level changes between two groups; **(D-E)** Disease gene network analysis for the significant pseudo-trajectory proteins. **(D)** X axis listed the enriched significant pseudo-trajectory proteins. The y-axis represents the disease enriched corresponding to the proteins listed at x - axis. For each disease (y axis), we used different color to lable the disease name. And in each enriched disease, we used same color as the disease name(y axis) by box shape to highlight the distinct enriched protein in each disease. **(E)**. X-axis was the EYO of enriched significant pseudo-trajectory proteins; y-axis was the −log10 estimate of significant trajectory proteins. The color of each curve corresponds to the color of the disease name in **(D).**

The top 30 hits included proteins known to be associated with AD, such as neurofilament (NEFH, NEFL), calcineurin complex (PPP3CA, PPP3R1), and 14-3-3 proteins (14-3-3 beta, gamma, zeta). Other proteins that have not been previously associated with AD, but were significant in our analyses, included the extracellular matrix binding (SMOC2, SLIT2), negative regulation of protein metabolic process (PEBP1, GPI, PTPA, CRKL, PIN1), cytoskeletal protein binding (PDLIM4, PDCD5, STMN2, TMOD3, MAPRE3, MAP1LC3A), cytosol function proteins (CHN1, DLG2),^52^ and other protein binding (TCEAL5, GARS1).^52,53^ However, other reported AD markers, including GFAP (p = 1.25×10^−05^) or TREM2 (p = 9.79×10^−03^), did not pass Bonferroni threshold **(Fig. S1A)**.

To determine the reliability of our pseudo-trajectory analysis, we also conducted an analysis based on mutation status as a phenotype, comparing carriers and non-carriers, regardless of clinical status (**Fig. 2B, Table S2)**. We identified 246 proteins to be associated with mutation status after Bonferroni correction **(Fig. S1B)**. We found that all proteins associated with the pseudo-trajectory (n=125) were also significant in this analysis (see supplementary materials). The top upregulated proteins associated with mutation status include CHIT1 (p= 7.97×10^−17^), NEFL (p=2.66×10^−18^), YWHAG (p=2.72×10^−26^), ITGA1/ITGB1 (p=1.18×10^−13^), and PPP3CA/PPP3R1 (p=1.10×10^−26^). There were two proteins that displayed downregulation in ADAD mutation carriers: NPTX2 (p = 1.61×10^−07^) and LRFN2 (p = 1.30×10^−06^; **Fig. S1C)**. When comparing the effect size of the proteins associated with both pseudo-trajectory and mutations status, we found strong correlation (R^2^ = 0.88, p = 9.79×10^−121^; **Fig. S1C)**, indicating that both methods captured the same overall findings, even though mutation status analysis seems to provide more statistical power, as 122 additional proteins passed Bonferroni threshold in this particular analysis (turquoise blue dots in **Fig.S1B, S1C**).

To confirm the reliability of our findings, we analyzed two other ADAD proteomic studies. ^15,16^ One study represents a technical replication as it used a different proteomics approach in the same patient cohort, and the other is a replication in an entirely independent dataset. Johnson et al., employed a Mass Spectrometry approach to measure 59 proteins in 286 mutation carriers and 184 noncarriers from DIAN.^16^ This study identified 33 proteins significantly associated with mutation status after correcting for EYO. There were 12 of our 125 pseudo-trajectory-significant proteins included on the Johnson study and all of them also significant and with consistent direction (**Table S3, Fig. S1D).** These proteins displayed a moderate effect size correlation, with an R² of 0.62 and a p-value of 2.30×10^−3^ **(Fig. S1D).** Of the remaining 21 proteins identified in Johnson’s study, 14 were present in our proteomic data and five exhibited nominal significance in our analysis **(Table S3)**. These 14 proteins presented an effect size correlation (R²) of 0.51 with a p-value of 4.30×10^−3^ **(Fig. S1D)**. Similar results were found when analyzing all proteins associated with mutation status in our analysis (See in the supplementary materials).

In the other ADAD proteomic study, Van de Ende et al. used Olink-based proteomics to measure 808 proteins in 22 MCs and 20 age- and sex-matched controls. They identified a total of 66 proteins passing significance after FDR correction.^15^ Of the 125 pseudo-trajectory significant proteins, 17 were present on the Olink panel **(Table S4)**. Of these 17 proteins, 15 proteins also passed FDR correction in Van de Ende et al.’s study. Among the remaining two, SFRP1 exhibited nominal significance, while SPP1 was not found significant in this analysis despite being significant in the Johnson et al.’s study. These 17 proteins showed a very high effect size correlation (R^2^ = 0.83, p = 4.54×10^−07^; **Fig. S1E)**. Regarding the other 51 significant proteins identified in van de Ende et al.’s study, 42 of them were also present in the SomaLogic panel, with 16 of them showing nominal significance in our analysis **(Table S4)**. These 42 proteins showed a low effect size correlation (R^2^ = 0.07, p = 0.10; **Fig. S1E)**. Similar results were found when analyzing all proteins associated with mutation status in our analysis (See in the supplementary materials).

When we integrated the data from both Johnson and van de Ende et al., out of the 125 pseudo-trajectories proteins, 24 unique proteins were present in at least one of these studies, and all of them exhibit nominal associations in one of those studies and consistent direction of association **(Table S5),** supporting the robustness of our findings. Similar results were found when comparing the 246 associated with mutation status. The results of these comparisons were included in the supplementary materials, supplementary figures **(Fig. S1F, S1G)** and Tables **(Table S6, S7)**.

### Comparison of significantly altered proteins in ADAD and sAD

We then, determined if the proteins associated with ADAD mutation status exhibit an association with sAD. This analysis aimed to provide additional validation for the proteins we initially identified in ADAD and identify potential differences and commonalities with sAD. To investigate this, we generated and analyzed proteomic data from the CSF samples obtained from 1,763 individuals diagnosed with sAD (A^+^T^+^: 848, A^−^T^−^: 915) from multiple cohorts (see in supplementary materials). Among the identified 125 significant pseudo-trajectory significant proteins, all of them were also significant after Bonferroni-correction in sAD with consistent direction of effect.

When looking at the 246 proteins associated with mutation status in DIAN, we found a very high correlation in the effect sizes between sAD and ADAD mutation status analyses (R^2^ = 0.67, p = 4.71×10^−63^, **Fig. S1H**). There were five proteins that were associated with sAD, also after Bonferroni correction, but in the opposite direction **(Table S8, Fig. S1H)**: DLK2 (ADAD p = 1.99×10^−07^, β =-0.025; sAD p = 2.77×10^−05^, β = 0.022), LRFN2 (ADAD p = 1.30×10^−06^, β = −0.054, sAD p = 3.34×10^−03^, β = 0.023), RELT (ADAD p = 2.27×10^−06^, β = −0.020, sAD p = 1.34×10^−03^, β = 0.016), GAGE2A (ADAD p = 4.37×10^−06^, β = −0.024, sAD p = 4.2×10^−03^, β = 0.018) and EPHA4 (ADAD p = 5.22×10^−06^, β = −0.030, sAD p = 1.08×10^−07^, β = 0.0569; **Fig. S1H**). DLK2, is implicated in neurite outgrowth,^54^ synaptic plasticity,^55^ and neuroprotection,^56,57^ and has been associated with anxiety and depression.^58^ EPHA4 has been reported to be involved in the progression of AD due to synaptic dysfunction, and enriched in neurons. GAGE2A has been widely studied in its immune system function in cancer, but its role in neurological disease remains unclear. However, the other two proteins seem to play a role in the blood-brain barrier (BBB). LRFN2 is an endothelial-specific protein that shows a significant decrease in protein levels in neurodegenerative diseases, including AD, Parkinson’s disease, and Lewy body dementia.^59^ Moreover, LRFN2 as one of GBA variants have been linked to dysregulation of glucocerebrosidase, an enzyme that has been reported to be dysregulated in Gaucher disease, which is one of the most common lysosome storage diseases. ^60^ RELT, a member of the tumor necrosis factor receptor superfamily, has been shown to contribute to the acquisition and development of barrier properties in the blood-brain barrier (BBB). Both TNFRSF21 and RELT (TNFRSF19) are downstream targets of the Wnt/beta-catenin signaling pathway in BBB endothelial cells. When the TNFRSF21/TNFRSF19 signaling is dysregulated, it can result in the breakdown of the BBB’s endothelial layer. It’s worth noting that the Wnt/beta-catenin signaling pathway is essential for central nervous system (CNS) angiogenesis but not for the development of peripheral vasculature. ^61,62^ These data highlight how the differential regulation of distinct proteins and alterations in endothelial cells and the BBB result in different disease outcomes (ADAD and sAD).

To investigate which proteins presented different effect sizes between sAD and ADAD, we identified proteins with effect sizes outside of the 95% CI for the regression for all the 246 proteins associated with ADAD. There were six proteins (CHIT1, SMOC1, SMOC2, NEFL, CAND1, PTPN11) that showed significantly higher effect size in ADAD compared to sAD **(Table S8, Fig. S1H)**. The protein with the highest difference in ADAD was CHIT1 (p = 7.97×10^−17^, β = 0.29). Chitotriosidase (CHIT1), a putative marker of microglial activation,^63^ has already been shown to be elevated in the CSF and peripheral blood of AD patients.^64^ Another protein with higher effect size in ADAD is CAND1 (p = 1.39×10^−14^, β = 0.09), which has been involved in modulating the ubiquitin-proteasome pathway (UPP).^65^ This pathway is pivotal in removing proteins, including the clearance of misfolded proteins, and plays a crucial role in controlling various cellular functions. CAND1 directly interacts with Cullins in their unneddylated form, regulating the assembly of Cullin-RING ligase complexes (CRLs). When Cullins are neddylated, CAND1 dissociates, allowing Cullins to form active CRLs, which are essential for targeting specific substrates for ubiquitination. Additionally, the Gain-of-Function mutation of PTPN11 (p = 4.32×10^−07^, β = 0.48) has been reported to associate the developing human brain, memory, and attention.^66^ Two other proteins with higher effect size in ADAD are SMOC1 and NEFL, that have already been proposed as AD biomarkers in previous studies.^15,67,68^ The only protein that exhibited a significantly lower effect size in ADAD was NPTX2, that is downregulated in both ADAD and sAD. NPTX2, known as a glutamate receptor, found in established synapses, has been implicated in non-apoptotic cell death of dopaminergic nerve cells **(Table S8, Fig. S1H)**.^69^

In summary, this analysis revealed a significant degree of overlap between ADAD and sAD, while also validating the mutation carrier proteomic analyses in sAD and highlighted several proteins and potential mechanisms that are different between sAD and ADAD.

### Earliest CSF proteomic changes

Due to the study design, that leverages mutation status and EYO, we estimated when proteins levels start changing in MCs compared to NCs in relationship of the EYO. Of the 125 identified proteins, 124 began to change before the estimated year of onset, with approximately 93% of these changes occurring within the range of −20 and −3 years (**Fig. 2C, Fig. S2)**. DOC2B was the only protein displaying changes one year after the estimated onset (change +1 yrs in relationship to EYO; **Table S9).** Established biomarkers changed between 11 to 17 years before onset: pTau (−17 yrs), total Tau (−12 yrs), and Aβ42 (−11 yrs). Among the 125 proteins significant in the pseudo-trajectory analyses, 12 initiated changes earlier than established biomarkers, such as, SMOC1 (−31 years), followed by SMOC2 at −28 years and PPP3R1, GAGE2D at −19 years (**Fig. 2A)**. Several proteins began to change around −18 years (SLIT2, PLCB1, TXNRD1) and −17 years (TMOD3, SPON1, MAPRE3, RABL6, PPP3CA, PPP3R1). Compared to the 33 significant proteins associated with EYO in Johnson et al., we reported 116 additional proteins with significant trajectories changes **(Table S1)**.

### Significant proteins in plasma suggest alterations in the presynaptic function

We employed an identical model to perform pseudo-trajectory and mutation status analysis between 325 MCs and 217 NCs from ADAD plasma samples. In the pseudo-trajectory analysis, out of 6,022 proteins, three proteins were significant after FDR correction (FDR p-value < 0.05; **Table S9, Fig. S3A**). Complexin-2 (CPLX2, p = 0.049) and Syntaxin 1A (STX1A, p = 0.049) were upregulated, while Vesicle Amine Transport 1 (VAT1, p = 0.049) was downregulated in MCs. In examining the time to symptom onset, CPLX2’s pseudo-trajectory began changing approximately seven years before symptom onset, whereas alterations in STX1A’s started around four years before symptom onset **(Fig. S3B)**.

In the mutation carriers vs. non-carriers analyses, nine proteins were associated with mutation status after FDR **(Fig. S3C, Table S10),** including CPLX2 and STX1A. Additional proteins identified in this analysis include MAD1L1, SMOC1, PPP4R3A, CPLX1, and CAST that were upregulated, while two proteins (SSBP1, VOPP1) were downregulated. SMOC1 was the only protein identified in both plasma and cerebrospinal fluid specimens **(Fig. S3D)**.

Overall, we identified a limited number of significant proteins associated with ADAD in plasma compared to CSF **(Fig. S3E, S3F)**. However, they revealed novel associations previously unreported in ADAD, highlighting differences from sAD. Pathway enrichment for the nine proteins of mutation carriers vs. non-carriers analyses point to alterations in the presynaptic function (see in the supplementary materials, **Fig. S4A, Table S11**).

### Dysregulated proteins are enriched in several known neurodegenerative diseases

We analyzed if the proteins identified in our analyses were reported to be implicated in other diseases by performing gene-disease network enrichment analysis. Of the 125 proteins identified on pseudo-trajectory analyses, 73 have been reported to be implicated on 55 different diseases **(Table S12)**. As expected, “Familial Alzheimer Disease (FAD)” was one of the top hits, with ten of the proteins being associated (MAP1LC3B, VSNL1, TPT1, PEBP1, NRGN, NEFL, HPRT1, ENO2, DLG4, ACHE; p = 6.28×10^−04^, log2 Fold Change (LogFC) =2.77, **Fig. S5A**).

The top ten enriched diseases predominantly were neurodegenerative and neurological disorders **(Table S12)**, such as “Creutzfeldt-Jakob disease” (p = 1.73×10^−05^, logFC =3.70), “Frontotemporal Dementia” (FTD, p = 2.26×10^−04^, logFC=2.74), and “senile plaques” (p = 5.20×10^−04^, logFC=2.83), involving a total of 40 proteins. Among these proteins, CHIT1, STMN2, NRGN, NEFL, GPI, ENO2, ACHE, YWHAZ, PIN1, GAP43, and PPIA served as critical connectors in the network of the 15 neurodegenerative diseases **(Fig. S5B)**. These key connectors play roles in various cellular functions such as postsynaptic signaling (NRGN, NEFL, GAP43, PIN1), neuron projection (STMN2, ENO2, GPI), and cell junction (PPIA, YWHAZ, ACHE),^52^ suggesting they may be common contributors across these diseases. We then investigated the overlapping proteins across the top ten diseases and observed that NEFL, ACHE, and GAP43 were the top three proteins recurrently enriched in these conditions (**Fig. 2D)**. These proteins are known to influence inflammatory responses, apoptosis, and oxidative stress.^70–73^ Moreover, GAP43 has been reported to be associated with rapid hippocampal atrophy, AD-signature hypometabolism, and AD-signature hypometabolism-related cognitive decline.^74^ Still, its link to ADAD was identified for the first time in our analysis. The unique proteins associated with each disease were also investigated (See the supplementary materials).^75–78^

Finally, we determined if the proteins enriched in specific diseases showed a distinct pattern in relation to the ADAD EYO (**Fig. 2E, Table S13)**. Mild cognitive disorder (MCD; p = 5.01×10^−03^, logFC=2.05) and neuropathy (p = 3.70×10^−03^, logFC=2.14) were characterized with significantly decreased NPTX2 (EYO - 10.09 yrs, β= −4.15×10^−03^, p = 1.34×10^−06^) protein level, and an increased NEFL (EYO −11.57 yrs, β=9.11×10^−03^, p=3.44×10^−19^) and ACHE (EYO −3.98 yrs, β=3.96×10^−03^, p=2.45×10^−07^) protein expression. On the other hand, proteins enriched in vascular dementia (DV, p = 5.01×10^−03^, logFC=2.74) and mild cognitive disorder (MCD, p = 5.01×10^−03^, logFC=2.05) showed the earlier EYO (< −15 yrs). These early changes were driven by TXNRD1 (EYO −17.82 yrs, β = 2.93×10^−03^, p = 4.05×10^−08^) and VSNL1 (EYO −15.72 yrs, β = 3.10×10^−03^, p = 3.18×10^−08^), followed by GAP43 (EYO −14.99 yrs, β = 4.45×10^−03^, p=1.35×10^−08^; **Fig. 2E)**. However, other disease groups such as FAD (p = 6.28×10^−04^, logFC =2.77) and FTD (p = 2.26×10^−04^, logFC=2.74) showed consistent effect sizes in all ranges of EYO and includes proteins such as NRGN (EYO −15.18 yrs, β=5.31×10^−03^, p = 3.24×10^−08^), SMURF1 (EYO −14.73 yrs, β=9.91×10^−04^, p = 2.14×10^−08^) and CHIT1 (EYO - 13.53, β=1.75×10^−02^, p = 7.30×10^−10^).

### Co-expression network analysis identified modules that capture the chronological progression in ADAD

Our analyses suggest that there are many pathways dysregulated in ADAD (**Table S14, Fig. S5C, S5D,** the overview of pathway enrichment for 125 proteins can be found in the supplementary materials), and their disruption may happen at different times during the disease curse. In order to disentangle this complexity, we performed co-expression network analyses and then pathway analyses in each module.

Co-expression network analysis identified four modules for the 125 significant pseudo-trajectory proteins: MEgrey (n=6 proteins), MEchocolate (M1, n=58), MEhotpink (M2, n=24), and MEdeepskyblue (M3, n=47; **Fig. S6A-S6C, Table S15, Fig. 3A)**. The MEgrey module comprised proteins not clustered into the other three modules, and therefore was not included for further analyses. We found that the nodes presented significantly different mean EYO, with M1 being enriched in proteins with the earliest changes (mean were EYO: −11.44 ± 5.92yrs), followed by M2 (−11.08 ± 3.52yrs), and M3 (−8.00 ±4.39 yrs**; Table S15, Fig. S6D)**. The difference between the mean EYO for M1 and M2 was not statistically significant, however, significant differences were observed when comparing the mean EYO of M1 and M3 (t-test: P =0.001) as well as M2 and M3 (t-test: P=0.004).

**Figure 3.**
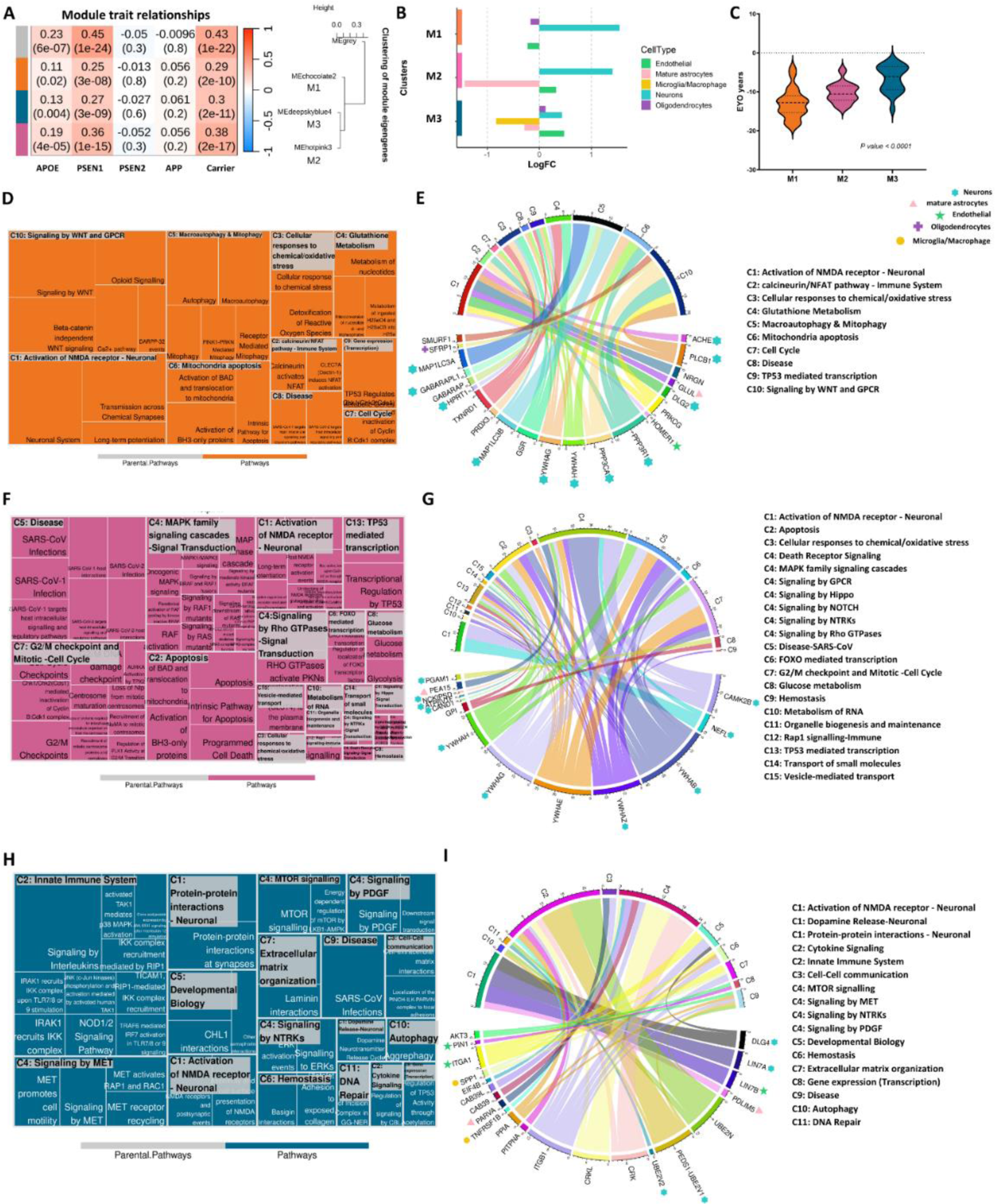
Co-expression network analysis of significant pseudo-trajectory proteins and pathway enrichment for each module. **(A)** Module-trait associations. Each row corresponds to a module eigengene, column to a trait. Each cell contains the corresponding correlation and P-value. The table is color-coded by correlation according to the color legend; **(B)** Cell type enrichment analysis for MEchocolate2 (M1), Mehotpink3 (M2) and Medeepskyblue4 (M3) clusters identified from WGCNA. The color bar showed on y-axis is consistent with the module color in **(A)**. Each cell type was assigned color as showed on the legend; **(C)** EYO comparison for functional identified proteins from Reactome pathway analysis. **(D-I)** Reactome pathway analysis for each module. Treemap **(D,F,H)** was used to present the significantly enriched pathways with summarized categories (C #, such as C1); chord diagram **(E,G,I)** were showing the enriched proteins in categorized pathways. The colored pattern labelled at proteins represent the different cell types and the colors were consistent with bar colors in cell type enrichment **(B)**.

The M1 module, which showed the earliest proteomic changes, was mainly enriched in neuronal-specific proteins (p= 1.07 ×10^−09^, logFC = 1.54) (**Fig. 3B, Table S16)**. Protein-protein interaction revealed enrichment in calcineurin complex (p =1.80×10^−02^, FC= 2.08), autophagosome membrane, p = 3.30×10^−03^, FC=1.44), depletion in postsynaptic density (p = 4.80×10^−03^, FC=0.89), and glutamatergic synapse (p = 5.20×10^−03^, FC=0.88) pathways **(Table S17, Fig. S7A)**. Of the 58 proteins present in M1, 21 proteins displayed significant enrichment in 27 different pathways. This subset of 21 proteins showed a mean EYO = −12.99±3.87 yrs) (**Fig 3C, Table S18)**. These 27 pathways were summarized into 10 super-pathways (the super-pathways are denoted as C1-C10, **Table S19**). The top pathway categories include NMDA receptor synapse signaling (C1: p =1.23×10^−02^, logFC=2.60), calcineurin/NFAT pathway (C2: p=1.53×10^−02^, logFC= 5.6), cellular stress (C3: p=1.71×10^−02^, logFC=4.47), Macroautophagy & Mitophagy (C5: p=1.53×10^−02^, logFC=5.63) and mitochondrial damage (C6: p=4.81×10^−03^, logFC=5.77; **Table S18, S19, Fig. 3D, 3E).** In these pathways, nine out of 21 proteins were present in the tier 1 or tier 2 of druggable genome database, such as proteins in the NMDA receptor synapse signaling (super-pathway C1; ACHE, PRKCG, GLUL), Glutathione Metabolism (C4: p=4.96×10^−02^, logFC= 3.05; GSR, TXNRD1, HPRT1), C6 (PPP3R1, YWHAG) and C10 (SFRP1; **Table S18)**. The mitochondrial damage pathway (C6) was driven by the 14-3-3 gamma (p=8.42×10^−08^, EYO −14.79 yrs) and 14-3-3 eta (p=6.21×10^−06^, EYO −10.65 yrs), which were some of the most significant associated proteins also with lowest EYO. The identified microtubule associated proteins (MAP1LC3A, MAP1LC3B) enriched in Macroautophagy & Mitophagy (C5, **Fig. 3D**) and the neuronal proteins (**Fig.3E, Table S18)** suggest that early neuron pathology changes may involve microtubule dysfunctions. Several proteins in this module are known to be part of the AD pathogenesis and not just mere biomarkers such as ACHE, HOMER1, ^79^ and calcineurin (PPP3R1/PPP3CA)^80^. Other proteins such as NRGN are known and validated AD biomarkers and also part of this module.^81^ In summary, this module seems to capture very early neuronal dysfunction due to the presence of ADAD mutations.

Similar to M1, proteins in M2 enriched in neuronal and endothelial cells (**Fig.3B, Table S18)**, but also with proteins that change trajectories after those from M1, suggesting the capture of later processes. Of the 24 proteins in M2, 13 were enriched in biological pathways (**Table S18-S19)**, and showed a mean EYO of −10.28 ± 3.87 years (**Fig. 3C)**. Pathway analyses revealed many processes related with apoptosis (C2: p=8.36×10^−02^, logFC= 7.65, **Fig. 3F, Table S19**), neuronal death (C4: p=2.96×10^−02^, logFC=6.66) and very early immune response (C12: p=2.96×10^−02^, logFC=6.24). Protein-protein interaction analysis revealed that 14-3-3 proteins (beta, epsilon, eta, gamma, zeta) interact with CAMK2B and neurofilament proteins (NEFH, NEFL) within the postsynaptic intermediate filament cytoskeleton (**Fig. S7B**). Unlike the other two modules, M2 is significantly marked by its involvement in inflammation signaling transduction pathways, and early immune responses. The eight identified proteins (the four 14-3-3 proteins, CAMK2B, NEFL, and PEA15) participated in and a part a complex biology pathway interaction (**Fig. 3G)**. The networks include MAPK family signaling cascades and signal transduction pathways involving G protein coupled receptors (GPCR), Hippo, Notch, neurotrophic tropomyosin-receptor kinase (NTRKs), and Rho GTPases (C4: p=5.41×10^−06^, logFC=4.99), as well as cell injury/death pathways, such as apoptosis (C2: p=8.36×10^−02^, logFC=7.65), and Rap1 signaling (C12: p=2.96×10^−02^, logFC=6.24).

The M3 module, which is closer to symptom onset, was enriched in microglia and astrocyte-specific proteins, along with pathways linked to the innate immune system (C2: p=4.35×10^−03^, logFC=2.62) and inflammation/infection (C9: p=4.00×10^−02^, logFC=2.14). Of the 47 proteins in M3, 20 were part of specific biological pathways, and showed an EYO of −6.65 ± 3.41 years (**Fig. 3C)**. Proteins enriched in microglia, such as CAB39/CAB39L, CRK/CRKL, EIF4B, and SPP1, were found to interact within signaling pathways of mammalian target of rapamycin (mTOR), MET,^82^ NTRKs,^83^ and Platelet-derived growth factor (PDGF),^84^ which are known to be involved in neurodegenerative disorders (**Fig. 3H)**. Additionally, ubiquitin proteins like UBE2N, UBE2V1, and UBE2V2, enriched in C2 (innate immune system, p=4.35×10^−03^, logFC=2.62) and C10 (Autophagy, p=4.71×10^−02^, logFC=4.08), were found to interact with the integrin alpha1-beta1 complex (ITGA1, ITGB1), involving intermediates like PPIA and CRK. This complex further interacts with the MPP7-DLG1-LIN7 complex (LIN7A, LIN7B) through DLG4 (**Fig. 3H, 3I, Fig. S7C).** Pathways related to cell-cell communication (C3: p=4.35×10^−03^, logFC=7.50), developmental biology (C5: p=9.71×10^−03^, logFC=6.34), and extracellular matrix organization (C7: p=2.22×10^−02^, logFC=3.68) appeared to reflect an effort to regain functionality by rebuilding and establishing new connections to maintain communication. PARVA, part of the cell-cell communication pathway, is involved in reorganizing the actin cytoskeleton and cell polarity, was identified for the first time in the context of ADAD. ^85,86^ In summary, by performing network and subsequent pathway analyses, we were able to provide a better overview of the pathways that are disrupted in ADAD at different stages of the disease.

### Predictive models of ADAD

We determined if the proteins identified in this study could predict mutation status (MCs vs NCs; independently of the clinical status). We used machine-learning approaches to identify the minimum number of proteins that maximizes the prediction power. We randomly split the CSF DIAN cohort into training (70%) and testing (30%) five times and used it for an iterative feature selection process. The LASSO model reduced the initial set of 125 proteins to nine key predictors that also showed a nominal or suggestive (p<0.1) association with ADAD in the multi-variate model (**Table S20**). The identified proteomic signature included some of the previously identified CSF AD-associated proteins, such as SMOC1, ^13,16^ calcineurin (PPP3CA/PPP3R1) or NPTX2, among others. ^87^ This model was validated in the testing set (30% of the DIAN CSF samples; n=143, MCs=94, NCs=49), showing strong prediction power for classifying ADAD mutation carriers, with an area under the curve (AUC) of 0.95 in the training set and 0.89 in the testing sets (**Fig. 4A, Table S21).** The AUC of this proteomic signature was significantly better than those for pTau181 (AUC: 0.80, p = 9.72×10^−03^), Aβ42 (AUC:0.72, p = 1.24×10^−02^) and pTau/Aβ42 (AUC: 0.80, p = 6.87×10^−06^) **(Table S21)**. Negative predictive values (NPV) were > 0.93 and positive predictive values (PPV) were > 0.70 in both training and testing and performing better than the classical AD biomarkers.

**Figure 4.**
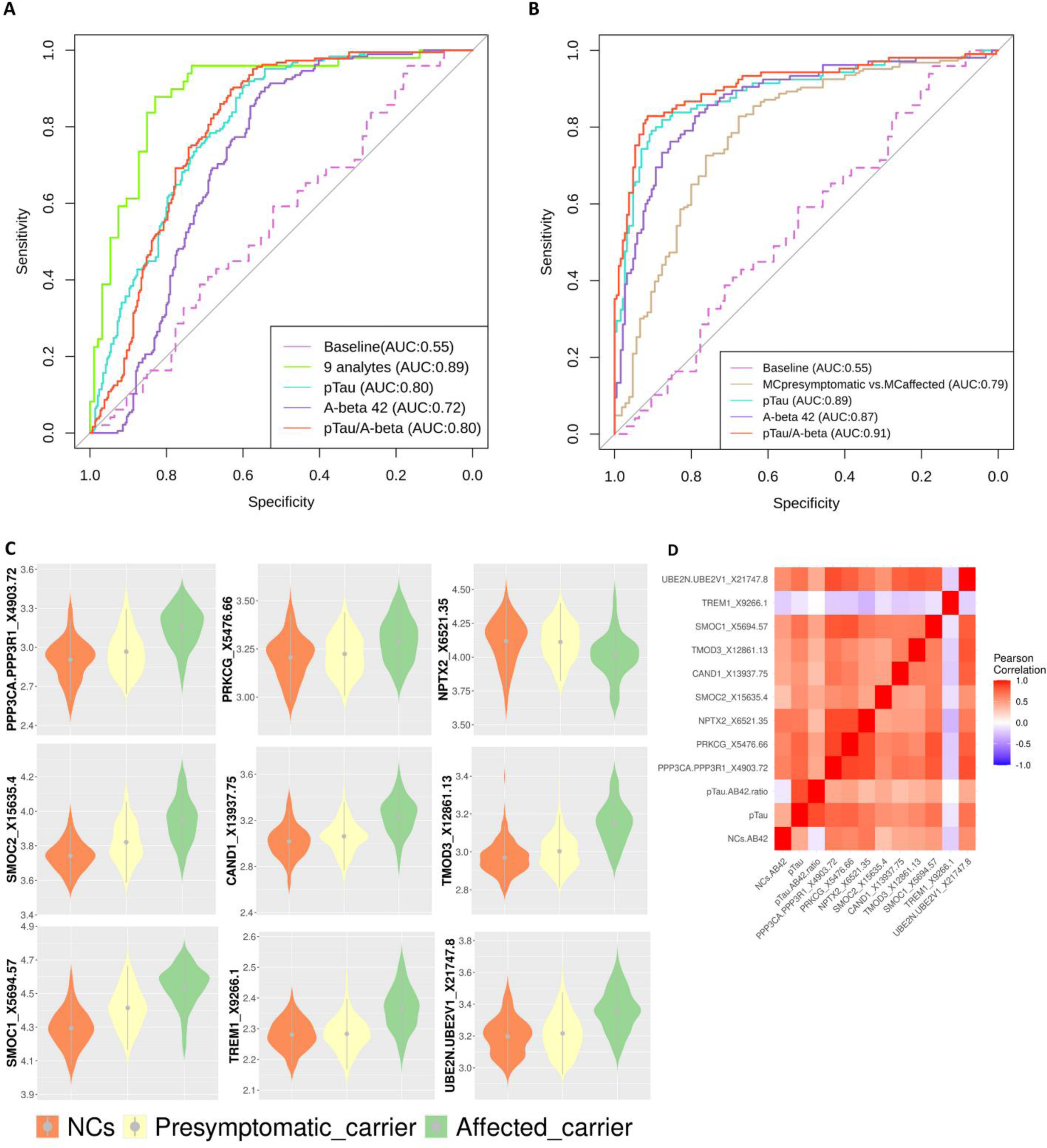
Prediciton models for ADAD. **(A)** The ROC curve for the testing dataset of nine proteins. **(B)** predictive model performance of nine proteins for presymptomatic carriers vs affected carriers. Aβ42, pTau, pTau/ Aβ42 prediction performance used as comparison **(A,B)**. **(C)** Levels of the nine proteins in NCs, pre-symptomatic carriers and symptomatic carriers. Grey dots with extended lines in each violin plot represent the median ±SD. **(D)** Pearson correlations for nine proteins in NCs.

Taking this analysis further, we categorized MCs into subgroups based on their clinical status at CSF draw date. Within the MCs there were 186 individuals without clinical symptoms (Pre-symptomatic) and 105 symptomatic individuals. We tested if this model could also distinguish the pre-symptomatic from the symptomatic individuals. We used the same proteomic signature (nine proteins) with the same weights and cut-offs values **(Table S20, S21)**. The nine proteins predictive model showed an AUC =0.79 (**Fig. 4B)**, the NPV (0.4) and the sensitivity (0.21; **Table S21**) were significantly lower than pTau181/Aβ42 (NPV 0.90 and sensitivity 0.83), suggesting that the nine proteins (**Fig. 4C,4D)** signature might be capturing very early disease changes.

## Discussion

In this study, we conducted an extensive analysis of CSF proteomics in the DIAN cohort (**Fig.1)**. Our goal was to uncover early changes in protein abundances, understand the underlying biology, and develop effective predictive models for the early diagnosis. We measured 6,163 proteins in 476 CSF samples and 6,022 proteins in 538 plasma samples from DIAN by Somascan® 7K Platform. We adopted a novel methodology, setting our work apart in the field of ADAD research. Firstly, we analyzed the CSF proteome changes in individuals with ADAD mutations, leveraging a high-throughput unbiased proteomic technology and a stringent pseudo-trajectory approach. In addition, our approach, unlike a previous study ^15^ that merely compared MCs to NCs, fully integrate the EYO to identify early proteomic changes. We broadened our analyses by including 1,763 CSF samples from sAD (6,163 proteins) from four independent cohorts, to serve as unbiased validation datasets.

We have identified 125 proteins (**Fig.2A, Table S1)** significantly different in pseudo-trajectories analysis and 246 significant proteins (**Fig.2B, Table S2)** associated with ADAD mutations status. These pseudo-trajectory changes could be traced back to 30 years before symptom onset (**Fig.2C, Table S1)**. For example, SMOC1 undergoes alterations as early as 15 years prior to established AD biomarkers like pTau, total Tau, and Aβ42. Some of the top findings include some known AD biomarkers such as NEFL and NRGN. However, most of the proteins identified in our analyses are novel findings, such as PPP3R1, which has been associated with disease progression and ptau levels in sAD,^80^ or GAGE2D that also showed protein changes 19 years before clinical onset. One recent study has reported high correlation between Somascan® and immunoassay measures (r > 0.9) ^34^, validating the robustness of platform used for proteomic profiling in this study.

We leveraged two previous studies^15,16^ to replicate our findings from the ADAD trajectory analysis. Of the 125 proteins identified in our analysis, 24 proteins were included in either of the previous studies **(Table S5)**, and all of them displayed significant associations in the same direction. Johnson et al., ^12^ represents a technical replication, as the same samples were analyzed to measure the levels of 33 proteins using Mass Spectrometry. The van der Ende et al. study^13^ represents a biological replication as 808 proteins were analyzed using Olink in 22 MCs and 20 controls not included in this study. These validation by mass spectrometry and Olink platforms, add confidence that the protein is reliably measured using different targeting methods.^15,16^ We also found that most of the proteins associated with different trajectories in MCs vs. NCs were also found to be associated with sporadic AD in a large dataset measured by Somascan 7K® (Table S8). These results support our findings and indicate a large overlap between ADAD and sAD. However, during these analyses, we identified five proteins (DLK2, LRFN2, RELT, GAGE2A, EPHA4) that exhibited opposite directions between ADAD and sAD. One potential explanation of this opposite effect could be due to the dynamic changes of these proteins in relation to disease status. In the pre-symptomatic phase of ADAD, some metabolic and synaptic plasticity markers (DLK2, LRFN2) ^55,88^ are observed to decreased in our analysis, possibly as a compensatory response to early pathological changes. A zebrafish study indicates that the loss of DLK2 is likely to impair neural stem cell function with long-term consequences for the telencephalon morphology.^89^ However, as ADAD progresses to the symptomatic phase, these markers might show an increase, indicative of deteriorating neuronal function and synaptic integrity. This hypothesis aligns with observations about heightened activation of EPHA4 signaling, which directly impacts motor neurons leading to their degeneration. ^90^ This pattern contrasts with the more linear progression seen in sAD, suggesting that the timing and progression of disease stages in ADAD vs. sAD could be a key factor in understanding these disparate protein change directions. Additionally, we found six proteins (CHIT1, SMOC1, NEFL, SMOC2, CAND1, PTPN11) with a higher effect size in ADAD. These findings suggest that there are distinct and diverse biological processes that are dysregulated in ADAD.

By analyzing a larger number of proteins than any of the previous studies and utilizing a novel approach to identify proteins with different trajectories, we identified 101 novel proteins with changes starting 22 years before onset, such as GAGE2D (EYO = −19.05 yrs) and PPP3R1 (EYO =-18.82 yrs). We then performed additional studies to determine the specific pathways that are implicated in disease. In order to disentangle the complex processes leading to disease, we first performed network analyses and then pathway enrichment analyses (**Fig. 2D, 2E, Table S12-S19)**. These analyses unveiled a significant enrichment within three distinct modules, each linked to different stages of the disease (**Fig. 5)**. In the earliest stage, the M1-module that capture early neuron pathology, displayed changes in cellular stress (GSR, PRDX3, TXNRD1), mitochondria damage (GABARAP, GABARAPL1, MAP1LC3A, MAP1LC3B, PPP3R1) and NMDA receptor synapse signaling (ACHE, PLCB1, NRGN, GLUL, DLG2, PRKCG, HOMER1). The proteins enriched in neurons indicated a depletion that might be caused by the cellular stress affecting their mitochondria and microtubules, leading to programmed cell death in these neurons. This transition and process was captured by first two modules (M1 and M2). As time progresses and the disease trajectory transitions to the M2-Signaling Transduction stage, these stimuli begin. Many studies have shown evidence of excitotoxicity of glutamatergic neurotransmission through postsynaptic NMDAR on the neurons.^91–93^ Some other studies suggested that various cellular stressors, such as oxidative stress and endoplasmic reticulum stress, could dysregulate MAPK pathways,^94,95^ which we observed in the M2 stage. These findings and evidence collectively contribute to our understanding and confirmation that oxidative stress or disruptions in glutamate metabolism, which initiate in the early and middle stages of ADAD, may promote the disease progression. Subsequently, under the regulation of cytokine, innate immune, and mTOR signaling pathways, microglia and macrophages become involved in an autophagy process that extends to other neurons and their microenvironments, as captured in the M3 module. However, the body consistently strives to maintain an equilibrium, attempting to repair and restore damaged neurons, axons, dendrites, and synapses. The presence of a dysfunction that exceeds the entire system’s compensatory capacity, gives rise to the onset of symptoms. This was mainly recapitulated in the M3 stage where dysregulation of ubiquitin-conjugating enzymes (UBE2N, PEDS1-UBE2V1) was observed. Non-degradative ubiquitin signaling is essential for homeostatic mechanisms crucial to neuronal function and survival^96^ and it plays a role in inflammatory processes,^96^ thus, supporting our hypothesis regarding self-rescue mechanisms at play during this stage.

**Figure 5.**
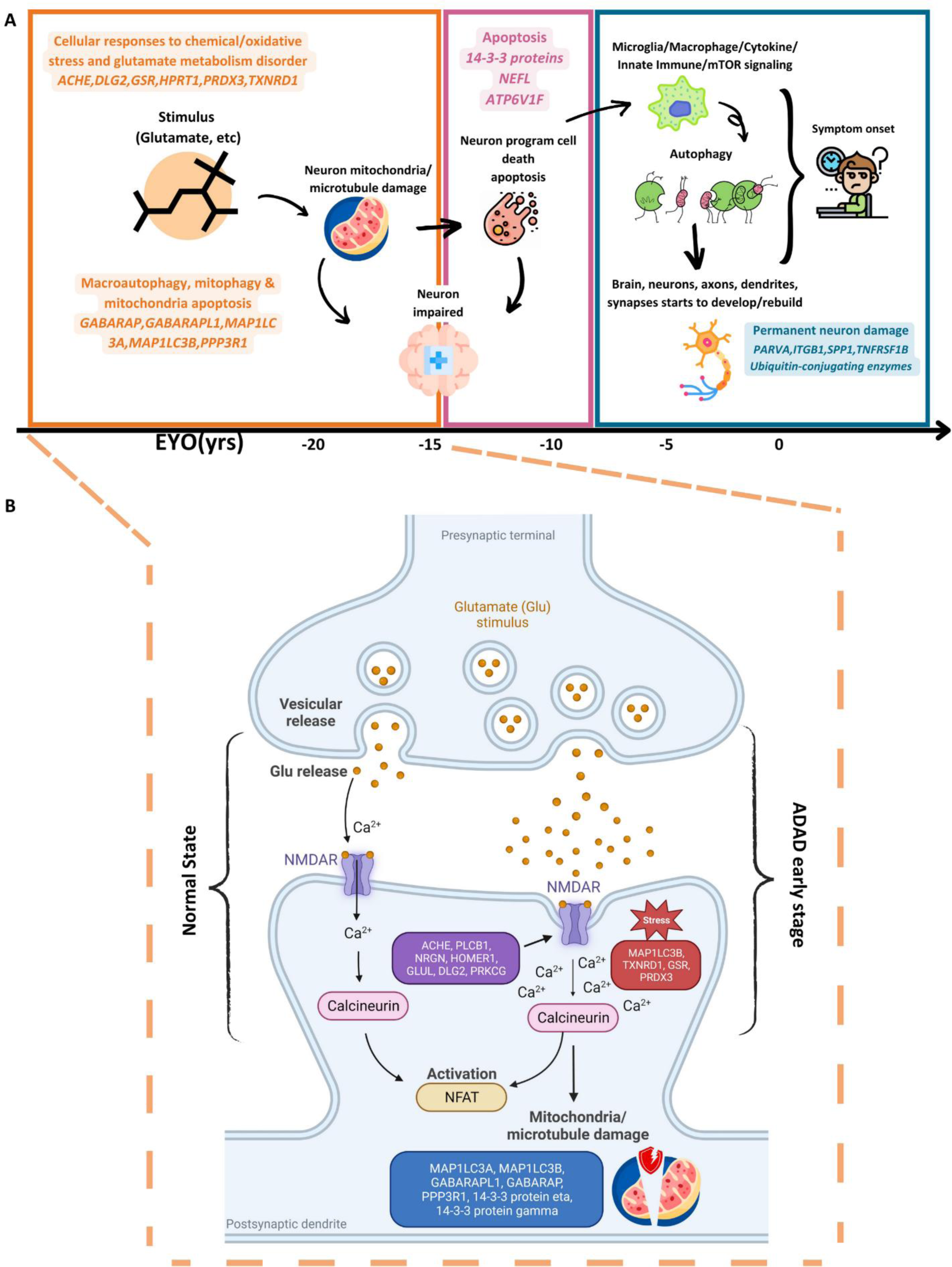
Multiple biological process trajectories summary. **(A)** Highlighted pathology process and enriched significant trajectory proteins in each module by chronological order. **(B)** Hightlighted biology process of early stage of ADAD (M1). It included normal state and early ADAD disease stage. X axis represents the EYO in years.

The proteins identified in this study were used to create a robust ADAD prediction model. We applied a LASSO model to narrow down the identified significant pseudo-trajectory proteins. The predictive model based on nine proteins accurately distinguished MCs from NCs, achieving a good performance with an AUC of 0.89 in the testing set, which is better than Aβ42, pTau and pTau/Aβ42 ratio (AUC < 0.80; p-value for AUC comparison p <0.012, **Fig. 4A, Table S21**). As some of the MCs were not symptomatic at the time of the lumbar puncture, we also analyzed if our model could distinguish between symptomatic and pre-symptomatic individuals carrying mutations. However, this model showed modest performance and low NPV (0.40) and poor performance compared to pTau/Aβ42 (p-value for AUC comparison p <0.026, **Fig.4B, Table S21**). As our protein discovery was focused on identifying proteins that showed the earlier changes in protein trajectories between MC and NC and not necessarily in clinical symptoms, it is reasonable to think that our model outperforms pTau/Aβ42 to identify MCs and not clinical symptoms. In the last decades, many studies have provided evidence for the prediction values of total Tau, pTau or pTau/Aβ42 for cognitive decline.^97–99^ Our model, while demonstrating lower NPV and sensitivity to distinguish mutation carriers with clinical symptoms against those without clinical symptoms compared to existing biomarkers, but the advantages of our model are quite obvious. They are mainly reflected in the following two aspects: 1) it is predominantly in detecting early protein changes, effectively distinguishing between early and late stages of ADAD. It achieves this with the highest PPV (0.87) and specificity (0.91) than the already known markers; 2) the nine proteins identified in our model can serve as secondary or confirmatory tests following initial screening with the known biomarkers that have high overall accuracy. This is particularly beneficial in situations where standard biomarkers are impractical, such as when patients cannot undergo imaging tests or when results are inconsistent with clinical symptoms. Overall, combining these nine proteins model with pTau/Aβ42 could have clinical utility as the nine proteins model could be used to identify individuals with very early changes and then pTau/Aβ42 could help to predict the age at onset.

Despite being the largest proteomic study in ADAD, and having sAD as comparison group, this study has several limitations. Firstly, our primary research cohort was the DIAN study, which, despite being unique, the largest ADAD cohort, and the most extensive in terms of proteomic screening, lacked a comparable cohort for systematic validation of our findings. Secondly, our pseudo-trajectory model relied on a multi-layers linear regression approach, even though certain pseudo-trajectory changes between MCs and NCs displayed non-linear patterns. Given the immense number of proteomic variables involved, testing and fitting each variable with the most appropriate model proved impractical. Thirdly, we clumped all MCs regardless of ADAD affected gene to compare with NCs during our analysis due to limited sample size. Therefore, we lack granularity in the understanding of the role of each of the particular mutations. Despite the limitations, our study offers valuable insights and warrants further investigation to provide a more comprehensive understanding of proteomic changes in ADAD.

In summary, our study leverages the DIAN participants and represents the most comprehensive proteomic analysis of ADAD to date. Our methodology employs extensive protein panels, covering a broad range of biological processes. The detection of numerous dysregulated proteins through a robust approach, exhibiting altered patterns early in the disease and maintaining statistical significance even after rigorous multiple testing corrections, highlights profound changes in the CSF proteome in ADAD. This reinforces the value of CSF proteomics in investigating the disease’s pathophysiology. Additionally, our study draws parallels between the CSF proteomes of ADAD and sAD, indicating the notable similarities, but also identifies differences that can lead to personalized medicine for those carrying any mutation in ADAD genes. Ultimately, our results lay the groundwork for creating new predictive models and identifying potential therapeutic targets, enhancing our understanding of ADAD and fostering the development of more effective future treatments.

## Supporting information

Main Figure 1 to Figure 5

Supplementary Methods and Results

## Acknowledgements

We thank all the participants and their families, as well as the many involved institutions and their staff.

This work was supported by grants from the National Institutes of Health (R01AG044546 (CC), RF1AG053303 (CC), RF1AG058501 (CC), U01AG058922 (CC), RF1AG074007 (YJS), the Chan Zuckerberg Initiative (CZI), the Alzheimer’s Association Zenith Fellows Award (ZEN-22-848604, awarded to CC), and Anonymous foundation.

The recruitment and clinical characterization of research participants at Washington University were supported by NIH P30AG066444 (JCM), P01AG03991 (JCM), and P01AG026276 (JCM).

This work was supported by access to equipment made possible by the Hope Center for Neurological Disorders, the NeuroGenomics and Informatics Center(NGI: https://neurogenomics.wustl.edu/) and the Departments of Neurology and Psychiatry at Washington University School of Medicine.

<u>DIAN resources:</u> Data collection and sharing for this project was supported by The Dominantly Inherited Alzheimer Network (DIAN, U19AG032438) funded by the National Institute on Aging (NIA), the Alzheimer’s Association (SG-20-690363-DIAN), the German Center for Neurodegenerative Diseases (DZNE), Raul Carrea Institute for Neurological Research (FLENI), Partial support by the Research and Development Grants for Dementia from Japan Agency for Medical Research and Development, AMED, the Korea Health Technology R&D Project through the Korea Health Industry Development Institute (KHIDI), Spanish Institute of Health Carlos III (ISCIII), Canadian Institutes of Health Research (CIHR), Canadian Consortium of Neurodegeneration and Aging, Brain Canada Foundation, and Fonds de Recherche du Québec Santé. This manuscript has been reviewed by DIAN Study investigators for scientific content and consistency of data interpretation with previous DIAN Study publications. We acknowledge the altruism of the participants and their families and contributions of the DIAN research and support staff at each of the participating sites for their contributions to this study.

FACE resources: The Genome Research @ Ace Alzheimer Center Barcelona project (GR@ACE) is supported by Grifols SA, Fundación bancaria ‘La Caixa’, Ace Alzheimer Center Barcelona and CIBERNED. Ace Alzheimer Center Barcelona is one of the participating centers of the Dementia Genetics Spanish Consortium (DEGESCO). Authors acknowledge the support of Instituto de Salud Carlos III (ISCIII), Acción Estratégica en Salud, integrated in the Spanish National R+D+I Plan and financed by ISCIII Subdirección General de Evaluación and the Fondo Europeo de Desarrollo Regional (FEDER “Una manera de hacer Europa”) grants PI13/02434, PI16/01861, PI17/01474, PI19/00335, PI19/01240, PI19/01301, PI22/01403, PI22/00258 and the ISCIII national grant PMP22/00022, funded by the European Union (NextGenerationEU).

## Data availability

The individual-level data from DIAN cannot be publicly shared. Due the rarity of this disease it makes all the data identifiable. This has been corroborated with the IRB and confirmed with the NIH, thus the data cannot be shared in public repositories. However, data is available to approved investigators via data request through https://dian.wustl.edu/our-research/for-investigators/diantu-investigator-resources/dian-tu-biospecimen-request-form/.

The proteomics and individual-level genetic data obtained from the ADNI cohort can be requested through ADNI’s website (adni.loni.usc.edu) after access has been approved.

The proteomics and individual-level genetic data obtained from the Knight-ADRC can be requested through the NIAGADS website (ng00130).

The proteomics and individual-level genetic data obtained from Fundació ACE Alzheimer Center Barcelona can be requested through Fundacio ACE’s website (www.fundacioace.com).

## Dominantly Inherited Alzheimer Network

James M. Noble, Gregory S. Day, Neill R. Graff-Radford, Jonathan Voglein, Ricardo Allegri, Patricio Chrem Mendez, Ezequiel Surace, Sarah B. Berman, Snezana Ikonomovic, Neelesh Nadkarni, Francisco Lopera, Laura Ramirez, David Aguillon, Yudy Leon, Claudia Ramos, Diana Alzate, Ana Baena, Natalia Londono, Sonia Moreno, Mathias Jucker, Christoph Laske, Elke Kuder-Buletta, Susanne Graber-Sultan, Oliver Preische, Anna Hofmann, Takeshi Ikeuchi, Kensaku Kasuga, Yoshiki Niimi, Kenji Ishii, Michio Senda, Raquel Sanchez-Valle, Pedro Rosa-Neto, Nick Fox, Dave Cash, Jae-Hong Lee, Jee Hoon Roh, Meghan Riddle, William Menard, Courtney Bodge, Mustafa Surti, Leonel Tadao Takada, Martin Farlow, Jasmeer P. Chhatwal, V. J. Sanchez-Gonzalez, Maribel Orozco-Barajas, Alison Goate, Alan Renton, Bianca Esposito, Celeste M. Karch, Jacob Marsh, Carlos Cruchaga, Victoria Fernandez, Brian A. Gordon, Anne M. Fagan, Gina Jerome, Elizabeth Herries, Jorge Llibre-Guerra, Allan I. Levey, Erik C. B. Johnson, Nicholas T. Seyfried, Peter R. Schofield, William Brooks, Jacob Bechara, Randall J. Bateman, Eric McDade, Jason Hassenstab, Richard J. Perrin, Erin Franklin, Tammie L. S. Benzinger, Allison Chen, Charles Chen, Shaney Flores, Nelly Friedrichsen, Nancy Hantler, Russ Hornbeck, Steve Jarman, Sarah Keefe, Deborah Koudelis, Parinaz Massoumzadeh, Austin McCullough, Nicole McKay, Joyce Nicklaus, Christine Pulizos, Qing Wang, Sheetal Mishall, Edita Sabaredzovic, Emily Deng, Madison Candela, Hunter Smith, Diana Hobbs, Jalen Scott, Johannes Levin, Chengjie Xiong, Peter Wang, Xiong Xu, Yan Li, Emily Gremminger, Yinjiao Ma, Ryan Bui, Ruijin Lu, Ralph Martins, Ana Luisa Sosa Ortiz, Alisha Daniels, Laura Courtney, Hiroshi Mori, Charlene Supnet-Bell, Jinbin Xu & John Ringman

