## Supplementary material for "Systematic proteomics in Autosomal dominant Alzheimer’s disease reveals decades-early changes of CSF proteins in neuronal death, and immune pathways": Main Figure 1 to Figure 5

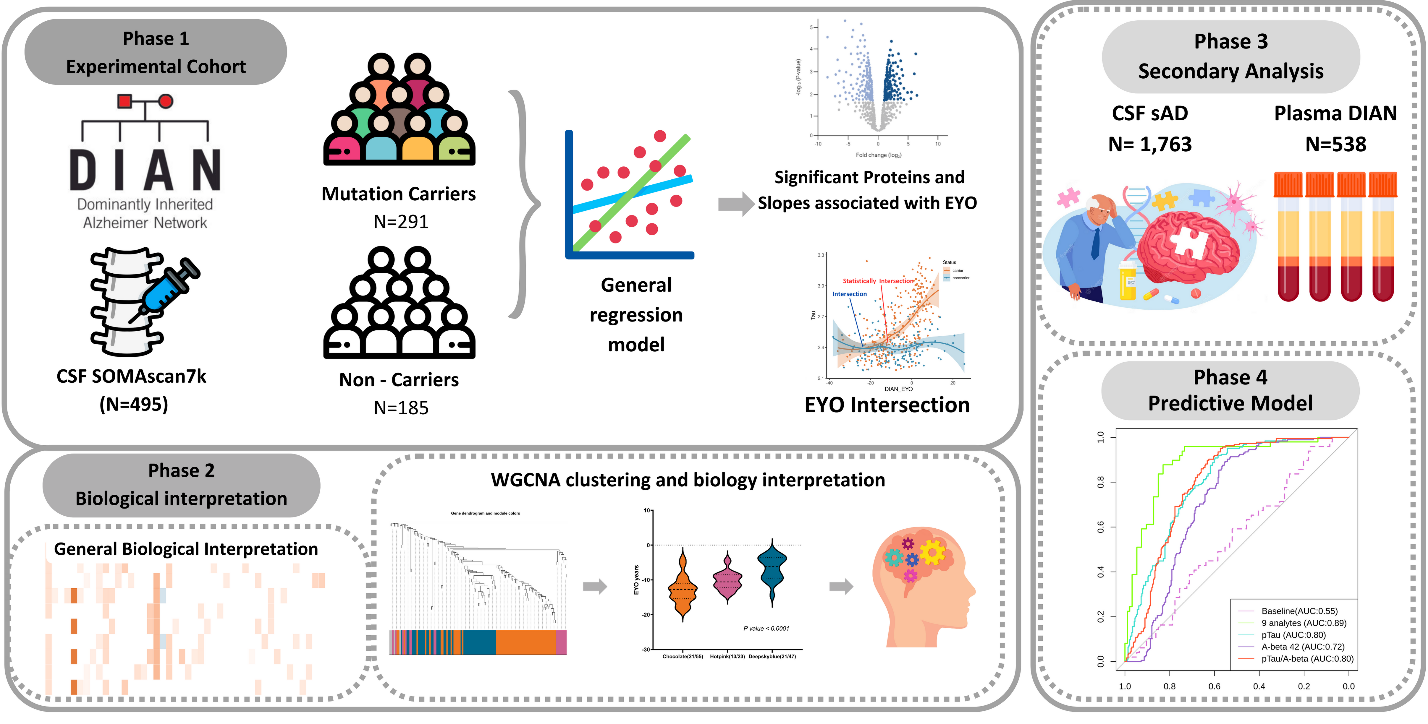

**Figure 1. Study overview.** In phase 1, proteins measured in the CSF sample were obtained with Somascan®, targeting 6,163 proteins from DIAN. In this stage, the experimental cohort contains 291 MCs and 185 NCs. Differential pseudo-trajectory analyses were performed between MCs and NCs. Trajectory intersections were calculated for significant pseudo-trajectory proteins. Biological functions were identified by protein co-expression network analysis and pathway enrichment.A total of 1,763 sAD CSF samples and 538 DIAN plasma samples were analyzed to validate the approach and contextualize the findings.Several publicly available external proteomic datasets were used to validate our findings as well (details in supplementary materials). Last, the LASSO model was used to select significant trajectory proteins and create predictive models for ADAD.

**Figure 2**

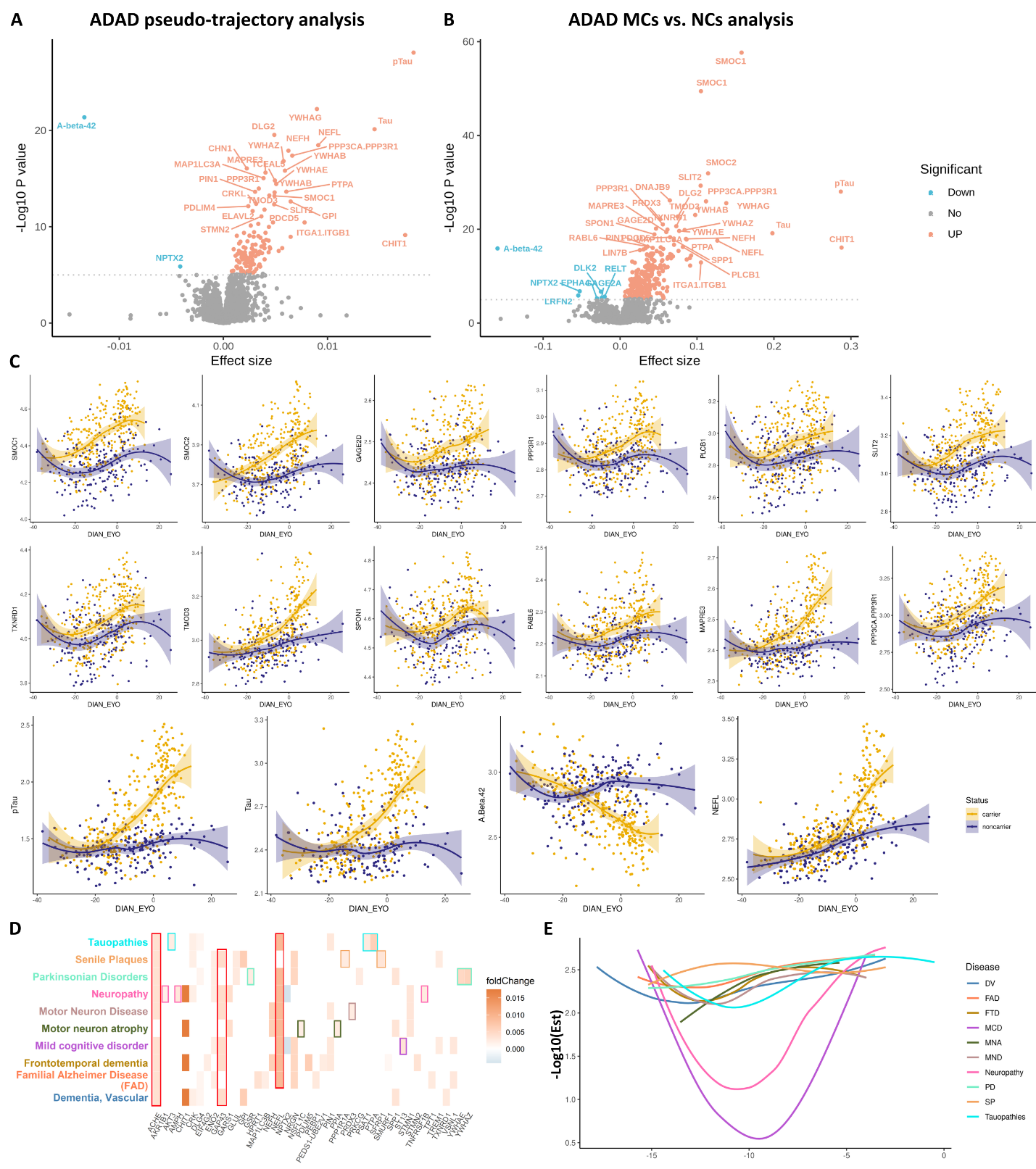

**Figure 2. Significant pseudo-trajectory proteins and significant proteins associated with ADAD mutation status in CSF**. (A-B) Volcano plots displaying the estimate change (x axis) against -log10 statistical differences (y axis) for all tested proteins. The red dots show the significantly upregulated proteins and the blue dots show the significantly downregulated proteins at Bonferroni threshold (p < 7.13×10⁻⁶).(A) Volcano plot for the pseudo-trajectory analyses comparing trajectories between MC and NC; (B) Volcano plot for the ADAD mutation status only; (C) Twelve significant pseudo-trajectory proteins that changed earlier than Tau, pTau, Aβ42. The ‘loess’ parameter in the plotting was applied to reflect the accuracy of the protein level changes between two groups; (D-E) Disease gene network analysis for the significant pseudo-trajectory proteins. (D) X axis listed the enriched significant pseudo-trajectory proteins. The y-axis represents the disease enriched corresponding to the proteins listed at x-axis. For each disease (y axis), we used different color to lable the disease name. And in each enriched disease, we used same color as the disease name(y axis) by box shape to highlight the distinct enriched protein in each disease. (E) . X-axis was the EYO of enriched significant pseudo-trajectory proteins; y-axis was the -log10 estimate of significant trajectory proteins. The color of each curve corresponds to the color of the disease name in (D)**.**

**Figure 3**

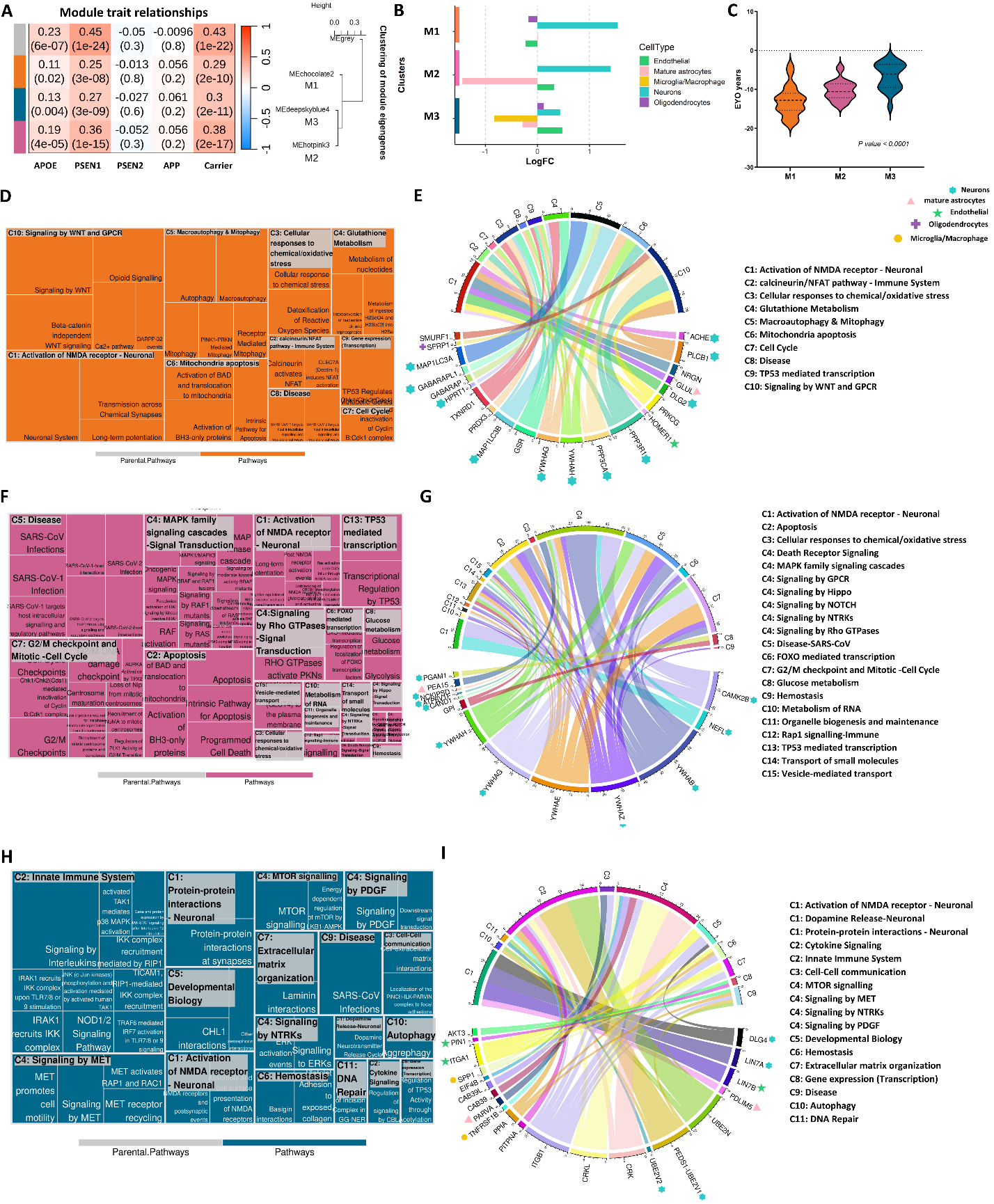

**Figure 3. Co-expression network analysis of significant pseudo-trajectory proteins and pathway enrichment for each module.** **(A)** Module-trait associations. Each row corresponds to a module eigengene, column to a trait. Each cell contains the corresponding correlation and P-value. The table is color-coded by correlation according to the color legend; **(B)** Cell type enrichment analysis for MEchocolate2 (M1), MEhotpink3 (M2) and MEdeepskyblue4 (M3) clusters identified from WGCNA. The color bar showed on y-axis is consistent the module color in **(A)**. Each cell type was assigned color as showed on the legend; **(C)** EYO comparison for functional identified proteins from Reactome pathway analysis. **(D-I)** Reactome pathway analysis for each module. Treemap **(D,F,H)** was used to present the significant enriched pathways with summarized categories (C #, such as C1); chord diagram**(E,G,I)** were showing the enriched proteins in categorized pathways. The colored pattern labelled at proteins represent the different cell types and the colors were consistent bar colors in cell type enrichment **(B).**

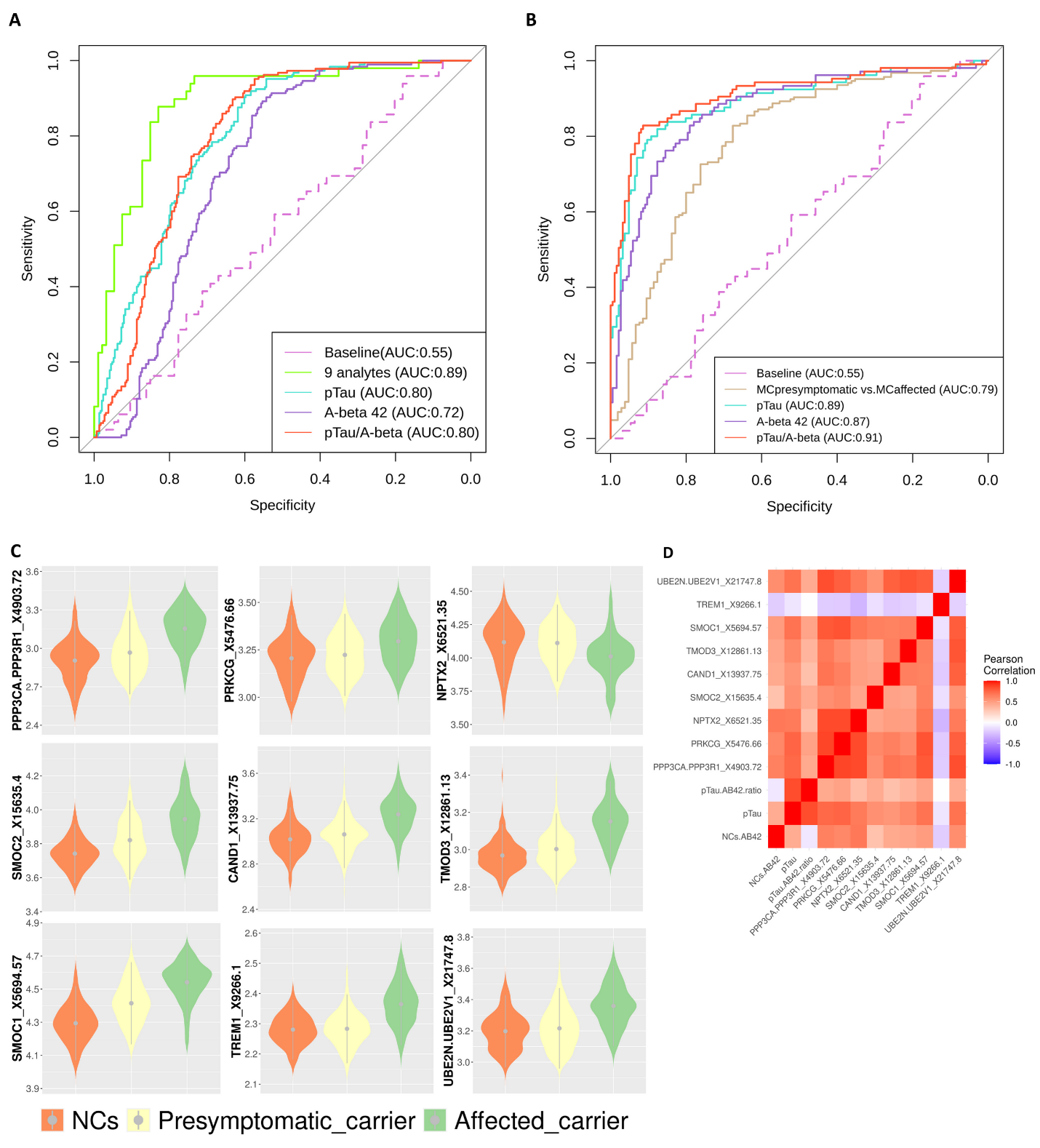
**Figure 4**

**Figure 4. Prediciton models for ADAD. (A)** The ROC curve for the testing dataset of nine proteins. **(B)** predictive model performance of nine proteins for presymptomatic carriers vs affected carriers. Aβ42, pTau, pTau/ Aβ42 prediction performance used as comparison **(A,B)**. **(C) L**evels of the nine proteins in NCs, pre-symptomatic carriers and symptomatic carriers. Grey dots with extended lines in each violin plot represent the median ±SD. **(D)** Pearson correlations for nine proteins in NCs.

**Figure 5**

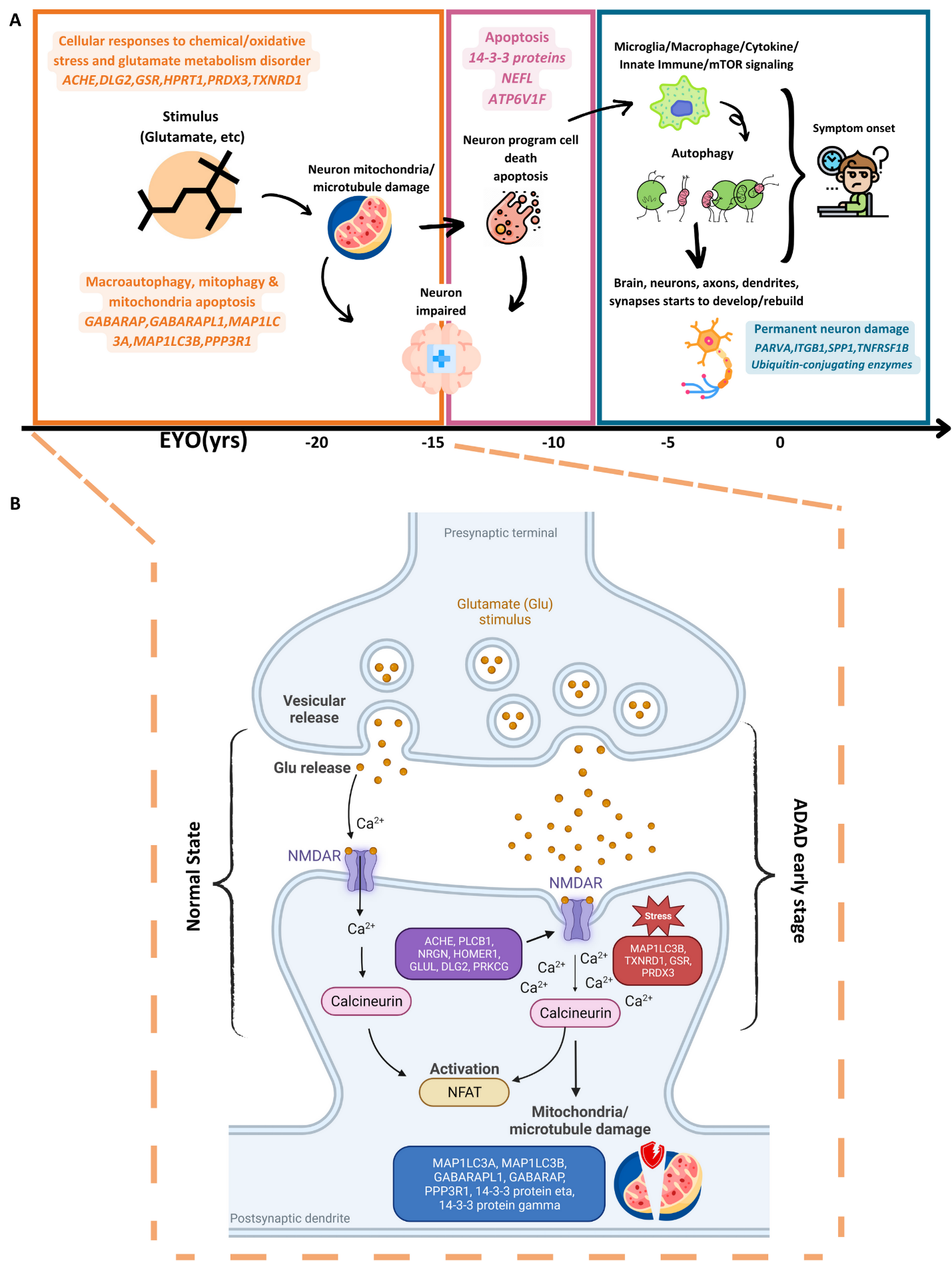

**Figure 5. Multiple biological process trajectories summary.** **(A)** Highlighted pathology process and enriched significant trajectory proteins in each module by chronological order. **(B)** Hightlighted biology process of early stage of ADAD (M1). It included normal state and early ADAD disease stage. X axis represents the EYO in years.
