## Supplementary Methods and Results for "Systematic proteomics in Autosomal dominant Alzheimer’s disease reveals decades-early changes of CSF proteins in neuronal death, and immune pathways"

**Extended Methods**

**Sporadic late-onset Alzheimer's Disease proteomics data cohorts**

In this study, we utilized the cerebrospinal fluid (CSF) proteomics data from four different cohorts, including Knight Alzheimer's Disease Research Center (Knight ADRC), the Alzheimer’s Disease Neuroimaging Initiative (ADNI), Ace Alzheimer Center Barcelona (FACE), Barcelona-1.

CSF samples used in this study were obtained from the Charles F. and Joanne Knight Alzheimer Disease Research Center (Knight ADRC, n=836) with 54.31% females, mean age 70.80 ± 8.51 year (yrs) old and 39% with at least one APOE ε4 allele; Alzheimer’s Disease Neuroimaging Initiative (ADNI, n=700) with 42.86% females, mean age 73.49 ± 7.50 yrs old and 50.43% with at least have one APOE ε4 allele; Fundació ACE Alzheimer Center Barcelona (FACE, n=618) with 59.22% females, mean age 72.14 ± 8.44 yrs old and 26.70% with at least have one APOE ε4 allele; and Barcelona-1 (n=132) cohorts with 54.55% females, mean age 68.16 ± 8.19 yrs old and 50.00% with at least have one APOE ε4 allele.

**Knight ADRC**

The Knight ADRC at Washington University School of Medicine has been recruiting and longitudinally assessing community-dwelling adults older than 45 years old since 1979. The Memory and Aging Project (MAP) at the Knight ADRC collects biofluids, and conducts annual clinical assessments, neuropsychological testing, neuroimaging studies, and autopsies of brain samples. Eligible participants may be asymptomatic or have mild dementia at the time of enrollment. All participants are required to participate in core study procedures, including annual longitudinal clinical assessments, neuropsychological testing, neuroimaging, and biofluid biomarker studies. Annual cognitive assessments of the participants were conducted by experienced clinicians. These assessments involved a semi-structured interview with both a knowledgeable collateral source and the individual displaying symptoms. The assessments followed the Uniform Data Set protocol of the National Alzheimer’s Coordinating Center and included a comprehensive neurological examination.^1^

Participants recruited in the Knight ADRC study are predominantly Non-Hispanic White individuals from North America (82.5%) and African-Americans (13.3%). So far, samples have been collected from a total of 5,510 participants, comprising 2,426 Alzheimer’s disease (AD) cases, 148 frontotemporal dementia (FTD) cases, 88 dementia Lewy body (DLB) cases, and 2,156 cognitively normal healthy individuals. Additionally, autopsy material is accessible for more than 1,182 participants, including 474 with fresh frozen parietal brain tissues ([https://dss.niagads.org/datasets/ng00127/](https://nam10.safelinks.protection.outlook.com/?url=https%3A%2F%2Fdss.niagads.org%2Fdatasets%2Fng00127%2F&data=05%7C01%7Ccruchagac%40wustl.edu%7C6a9d0b6406594f932f2908db2818eb8c%7C4ccca3b571cd4e6d974b4d9beb96c6d6%7C0%7C0%7C638147859699042664%7CUnknown%7CTWFpbGZsb3d8eyJWIjoiMC4wLjAwMDAiLCJQIjoiV2luMzIiLCJBTiI6Ik1haWwiLCJXVCI6Mn0%3D%7C3000%7C%7C%7C&sdata=NWgb32prewnfroBlyJ%2FgO9KeOb8tADNyAJd%2BUguFe%2Fk%3D&reserved=0)). Multi-tissue data from brain, CSF, and plasma has been utilized for generating multi-omics data encompassing genetics, epigenomics, transcriptomics, proteomics, metabolomics, and lipidomics with the aim of identifying new risk and protective variants for dementia as well as novel potential drug targets. Participants in the Knight ADRC were enrolled only if they exhibited cognitive normalcy, with a global clinical dementia rating® (CDR®) score of 0 at the time of enrollment. Study clinicians evaluate and make clinical diagnoses of incident dementia at the conclusion of each annual assessment. These diagnoses are formed by integrating results from both the clinical assessment and bedside measures of cognitive function.^2^ Dementia diagnoses followed the criteria established by the National Institute of Neurological Disorders and Stroke and the National Institute on Aging-Alzheimer's Association (NIA-AA) Work Group criteria for participants assessed after 2011.^3,4^ Diagnoses of AD dementia adhered to criteria developed by working groups from the NIA-AA.^4^ Additionally, diagnoses of vascular dementia conformed to the NINDS-AIREN criteria.^5^ Additional information about the Knight ADRC cohort is available at their website (knightadrc.wustl.edu).

**ADNI**

CSF proteomics data used in the preparation of this manuscript was obtained from the ADNI database (<https://adni.loni.usc.edu/>). Launched in 2003, ADNI represents a public-private partnership led by Principal Investigator Michael W. Weiner, MD. The primary objective of ADNI study has been to investigate whether the combination of serial magnetic resonance imaging (MRI), positron emission tomography (PET), other biological markers, and clinical and neuropsychological assessments can effectively measure the progression of mild cognitive impairment (MCI) and early AD.

**FACE**

We also acquired CSF samples from FACE, a private non-profit organization dedicated to AD research. Established in 1995 and based in Barcelona, FACE has diagnosed over 30,000 patients, collected 20,000 blood samples, 1,831 cerebrospinal fluid samples, and analyzed nearly 13,000 genetic samples.^6,7^ Additionally, it has been involved in nearly 150 clinical trials during its existence. Additional information about the FACE cohort is available at their website ([www.fundacioace.com/en](http://www.fundacioace.com/en)).

**Barcelona-1**

For this study, CSF samples were also sourced from Barcelona-1, a study led by the University Hospital Mutua de Terrassa in Terrassa, Spain. Barcelona-1 is a longitudinal study comprising approximately 300 individuals. Only those individuals, diagnosed with MCI or more severe conditions, underwent PET scans and CSF collection and follow-up analyses were conducted to monitor disease progression. The study encompassed individuals with diagnoses of subjective memory complaints (SMC), MCI, AD dementia (ADD), and non-AD dementias (non-ADD).

**Demographic information of sAD participants at the time of the CSF draw**

| Cohort | Knight ADRC | FACE | ADNI | Barcelona-1 |
| --- | --- | --- | --- | --- |
| Sample size | 836 | 618 | 700 | 132 |
| Males (%) | 45.69 | 40.8 | 57.1 | 45.45 |
| Age (mean) | 70.8 | 72.1 | 73.5 | 68.16 |
| Age (SD) | 8.51 | 8.44 | 7.5 | 8.19 |
| A+T+ (%) | 20.45 | 51.6 | 40 | 59.09 |
| A+T- (%) | 55.62 | 34.8 | 31.4 | 11.36 |
| A-T- (%) | 23.92 | 13.6 | 28.6 | 29.55 |
| APOE4+ (%) | 39 | 26.7 | 50.4 | 50 |

**ATN Classification**

To ascertain amyloid/tau classification for each sample, we relied on AD-specific biomarkers, namely amyloid beta 42 (Aβ42) and phosphorylated tau-181 (pTau), both measured in the CSF.^8^ For the classification of each sample based on amyloid and tau positivity, we conducted dichotomization utilizing the *mclust()* R package.^9^ The process of dichotomization was executed independently for each cohort (Knight ADRC, ADNI, FACE, and Barcelona-1). The levels of Aβ42 and pTau were log10-transformed to approximate a normal distribution. They were further normalized using a z-score transformation, which ensured they had a mean of 0 and a standard deviation (SD) of 1. Outliers were identified and subsequently removed based on a cutoff of 3 times the SD from the mean. Following the removal of outliers, the standardization by z-score was recalculated for the data. In case of Stanford ADRC cohort, we utilized the AT classification status provided by the study, which was also measured in the CSF.

**Knight ADRC**

In the case of the Knight ADRC cohort, measurements for both Aβ42 and pTau were conducted using the LumiPulse® G platform by Fujirebio US, Inc., based in Malvern, PA. However, for seventeen samples with missing LumiPulse® values, we utilized Innotest (Fujirebio) values as an alternative. This decision was informed by the high correlation observed between the two platforms, with a correlation coefficient (R^2^) of 0.73 for Aβ42 and 0.86 for pTau. Dichotomization was carried out for a total of 948 subjects for CSF Aβ42 and 944 subjects for CSF pTau. A specific cutoff of z-score = -0.20 was determined for Aβ42, which corresponded to a raw value of 630 pg/mL. For Aβ42, samples with values below 630 were classified as Aβ42-positive (A^+^). As for pTau, a cutoff of z-score = 0.61 was determined, which corresponded to a raw value of 62.9. Samples with values above 62.9 were considered pTau-positive (T^+^).

**ADNI**

In the ADNI cohort, Aβ42 measurements were conducted using Innotest by Fujirebio, while pTau measurements were performed using Elecsys by F. Hoffmann-La Roche Ltd in Switzerland. Dichotomization was carried out for 749 subjects based on Aβ42 measurements and for 745 subjects based on pTau measurements. For Aβ42, a z-score cutoff of 0.616 was determined, corresponding to a raw value of 196 pg/mL. Samples with values below 196 were classified as Aβ42-positive (A^+^). For pTau, a z-score cutoff of 0.197 was identified, corresponding to a raw value of 27.8. Samples with values above 27.8 were considered pTau-positive (T^+^).

**FACE**

In the FACE cohort,^10^ both Aβ42 and pTau measurements were conducted using Innotest (Fujirebio). This analysis included 632 samples. For Aβ42, a z-score cutoff of 0.468 was established, which corresponds to a raw Aβ42 value of 856 pg/mL. Samples with values below 856 were categorized as Aβ42-positive (A^+^). For pTau, a z-score cutoff of -0.018 was identified, corresponding to a raw value of 67. Samples with values greater than 67 were classified as pTau-positive (T^+^).

**Barcelona-1**

In the Barcelona-1 cohort, both Aβ42 and pTau measurements were performed using Innotest (Fujirebio). This analysis involved 231 samples. For Aβ42, a z-score cutoff of 1.04 was determined, which corresponds to a raw Aβ42 value of 1325 pg/mL. Samples with values below 1325 pg/mL were classified as Aβ42-positive (A^+^). For pTau, a z-score cutoff of -0.163 was identified, corresponding to a raw value of 58. Samples with values above 58 were categorized as pTau-positive (T^+^).

**Differential pseudo-trajectory analysis**

In the first step, we employed a linear regression, *lm()* function from *stats* package,^11^ to build the model with log 10-transformed protein abundance as the dependent variable, DIAN-EYO as the independent variable, sex, and SV as covariates for MCs and NCs, separately (model 1)

Formula 1:

Fit_MC_: log10 (protein level) ~ DIAN_EYO + sex + sv2 + sv1

Fit_NC_: log10 (protein level) ~ DIAN_EYO + sex + sv2 + sv1

Next, we obtained the coefficient differences of EYO generated from both status as slope differences:

Formula 2:

δSlope= coef(Fit_MC_)["DIAN_EYO"] - coef(Fit_NC_)["DIAN_EYO"]

Futher, we calculated the standard error (SE):

Formula 3:

SE= $\sqrt{SE_{MC}^{2}+SE_{NC}^{2}}$

Lastly, we calculated P-Value of differential abundance of proteins associated with EYO by pnorm() function and set lower.tail by FALSE:

Formula 4:

*P-Value* = 2* *pnorm*(|abs ($\frac{\delta\mathrm{Slope}}{SE}$)|)

**Pseudo-trajectory intersections**

Different from regular intersections of two curves, we calculated pseudo-trajectory intersections by following steps (model 2):

We firstly used *predict()* function to obtain the predicted 95% confidence interval (CI) of input protein value from Formula 1 for building a new linear regression models.

Formula 5:

Predict_MC_ = *predict*(Fit_MC_, interval=”prediction”)

Predict_NC_ = *predict*(Fit_NC_, interval=”prediction”)

Next, we retrieved lower bound (lwr) and upper bound (upr) of predicted 95% CI of each status and merge with the original matrix of each status to form a new matrix. Then we used either predicted lower bound or upr of specific protein levels from each new matrix to build a new linear regression model. Each time we choose lower bound or upr depending on the estimate of each protein. For example, if estimate of a protein is > 0, then we choose lower bound from MCs matrix and upr from NCs, vice versa.

Formula 6:

Fit_MC__new: _log10_(protein level)_lwr/upr_~ DIAN_EYO + sex + sv2 + sv1

Fit_NC__new: _log10_(protein level)_lwr/upr_ ~ DIAN_EYO + sex + sv2 + sv1

Then we obtained the two coefficients from the new linear regression models (Formula 6), and applied *solve()* function to calculate the pseudo-trajectory intersections for each protein.

**Differential protein level analysis associated with ADAD mutations**

Differential protein level across different status (MCs vs. NCs) in DIAN cohort and (A^+^T^+^ vs. A^-^T^-^) in sporadic late onset AD cohorts were detected using the linear regression model. We used log 10-transformed protein level as the dependent variable, status as the independent variable, where age at CSF draw, sex, and first two SV were used as covariates (Formula 7).

Formula 7:

_log10_(protein level) ~ Status + age at draw date + sex + SV1 + SV2

**Extended Results**

**Significant protein associated with ADAD mutation status**

We performed association analyses for mutation status including age, sex and SVs as covariates. We identified 246 proteins that passed Bonferroni-correction **(Fig. 2B, Table S2)** and 1,083 proteins nominally significant (p < 0.05). Among the 246 significant proteins, six proteins were downregulated in MCs (NPTX2, LRFN2, EPHA4, DLK2, GAGE2A, and RELT). The remaining 239 proteins were upregulated in MCs **(Fig. 2B)**. Of the 125 proteins that were significant after Bonferroni in the pseudo-trajectory analyses, 124 proteins also passed Bonferroni for mutation status and the rest were at least nominally significant.

**Replicates for significant analysis proteins associated with ADAD mutations status**

We utilized Johnson and van de Ende’s studies again to exam the consistency and significance of the proteins associated with ADAD, as identified through our mutation status analysis. In the case of the Johnson et al. study, which assessed 59 proteins, 15 of the 246 ADAD-associated significant proteins from our study were included, and all of them proved to be significant as well **(Table S6)**. These 15 proteins exhibited a notably high effect size correlation (R^2^ = 0.78, p = 1.80×10^-05^, **Fig. S1F**). Among the remaining 18 proteins in Johnson et al.'s study, 12 were present in our investigation, and eight of them displayed nominal significance in our results. But the effect size correlation between these 12 proteins in Johnson et al.'s study and our study were comparatively lower (R^2^ = 0.38, p = 0.03, **Fig. S1F**).

When comparing our findings to those of van de Ende's study, we showed that out of the 246 ADAD-associated significant proteins in our study, 33 proteins were also detected on the Olink panel used in van de Ende's research **(Table S7)**. Of these, 23 proteins passed FDR correction in van de Ende’s study, indicating a strong effect size correlation (R^2^ = 0.62, p = 5.68×10^-^^8^, **Fig. S1G).** Seven proteins exhibited nominal significance (TFPI, SNCG, THBS4, FIS1, CRIM1, FLT1, SFRP1), while three proteins (LRP1, RELT, SPP1) did not demonstrate any significance in van de Ende's study. Within the remaining 43 significant proteins identified in van de Ende's study, 34 proteins were also observed in our investigation, and out of those, ten proteins showed nominal significance in our ADAD mutation status results. However, the 34 proteins common to both studies displayed a considerably lower effect size correlation (R^2^ = 0.03, p = 0.363, **Fig. S1G)**

**Significant proteins in plasma suggest alterations in the presynaptic function**

To better understand the biological implications, we conducted pathway enrichment analysis on the significant proteins identified in the plasma mutation status analysis. In this analysis, two proteins (STX1A, CPLX1) were enriched in “Neurotransmitter release cycle” pathway (FDR p-value = 3.41×10^-04^, **Fig. S4A, Table S11).** This pathway included neurotransmitters such as glutamate, acetylcholine, and serotonin. In addition, CPLX2 works with CPLX1 to maintain a balance between synchronous and asynchronous neurotransmitter release, which is crucial for proper synaptic function and information processing.^12^ Notably, CAST was enriched in “Deregulated CDK5 triggers multiple neurodegenerative pathways in Alzheimer's disease models” (FDR p-value = 3.21×10^-02^), a pathway belonging to a group of programmed cell death diseases. Some other important proteins that are significantly altered in ADAD plasma but not part of already known biological pathways include VOPP1, PPP4R3A, SSBP1, and MAD1L1, among others. VOPP1, a known oncogene, whose upregulation is associated with cancer cell survival and proliferation,^13^ was downregulated in ADAD plasma samples. This is in line with recent studies reporting the inverse correlation between AD and cancer.^14,15^ In a recent GWAS study, PPP4R3A was reported to reduce the risk of AD progression to dementia,^16^ possibly due to its role in synaptic plasticity.^17^ SSBP1 is a protein that is crucial in DNA replication, repair, and maintenance.^18^ It has been reported that its mutation could impair mitochondrial DNA and lead to nephropathy and reduced nerve size.^19^ MAD1L1 is part of the spindle assembly checkpoint, which ensures accurate chromosome segregation during mitosis.^20^ In studies related to psychosis-related diseases, alterations in MAD1L1 methylation and transcription have been shown to mediate the risk for schizophrenia.

Furthermore, we compared the significant proteins identified from the CSF and plasma analysis in ADAD. Only four proteins were significant in both tissues, STX1A, CPLX2, SMOC1, and CPLX1, suggesting low overlap across these tissues. In fact, we did not observe a strong correlation of effect size across significant proteins in the pseudo-trajectory (R^2^=0.03, p =0.16) and mutation status (R^2^=0.01, p =0.06).

**CSF ADAD-associated proteins suggest overlap with other neurodegenerative disease**

We analyzed if the proteins associated with ADAD were also associated with other diseases. Mutations in HPRT1 can lead to Lesch-Nyhan syndrome, characterized by severe neurological disorder with early onset.^21,22^ In addition, STMN2, a member of the stathmin family, is implicated in microtubule dynamics and was uniquely enriched in FTD in our analysis.^23^ Likewise, mutation of NSFL1C has been associated with the inclusion body myopathy in motor neuron disease (MND) such as amyotrophic Lateral Sclerosis (ALS).^24^ These examples suggested that ADAD may share some similarities with other neurological diseases in terms of pathological processes and symptoms.

**Significant pseudo-trajectory proteins pathway enrichment**

We also conducted a pathway enrichment analysis to gain insights into the biological functions of the 125 significant pseudo-trajectory proteins. There were 59 pathways significantly enriched for the 125 identified proteins **(Table S14)**.

The most overrepresented pathway is the “Neuronal System”, (FDR p =7.80×10^-04^, logFC =1.96; **Fig. S5C),** included 13 proteins, cholinergic synapse proteins (ACHE, CAMK2B, PLCB1, PRKCG), glutamatergic synapse proteins (GLUL, HOMER1), signaling proteins (DLG2, DLG4), cell communication proteins (LIN7A, LIN7B, NRGN), proteins implicated on anatomical structure development (PDLIM5) and known AD biomarkers (NEFL).^25^ And these 13 proteins played as key connectors in pathways involved with NMDA receptor associated activities **(Fig. S5D)**, such as “Assembly and cell surface presentation of NMDA receptors” (FDR p =5.78×10^-05^, logFC =4.16). Another significant pathway was “Activation of BAD and translocation to mitochondria” (FDR p =6.01×10^-09^, logFC = 5.83) and belongs to the super-pathway “Intrinsic Pathway for Apoptosis” (FDR p = 2.08×10^-05^, logFC = 3.69, **Fig. S5D**). This super-pathway is connected with “Autophagy” (FDR p = 1.78×10^-02^, logFC =2.28, **Table S14, Fig. S5D**), through the 14-3-3-proteins (14-3-3 beta, epsilon, gamma, zeta) proteins, which are likely capturing early neuronal death and autophagy events in ADAD.
