## Supplementary Figure 1 to Figure 7 for "Systematic proteomics in Autosomal dominant Alzheimer’s disease reveals decades-early changes of CSF proteins in neuronal death, and immune pathways"

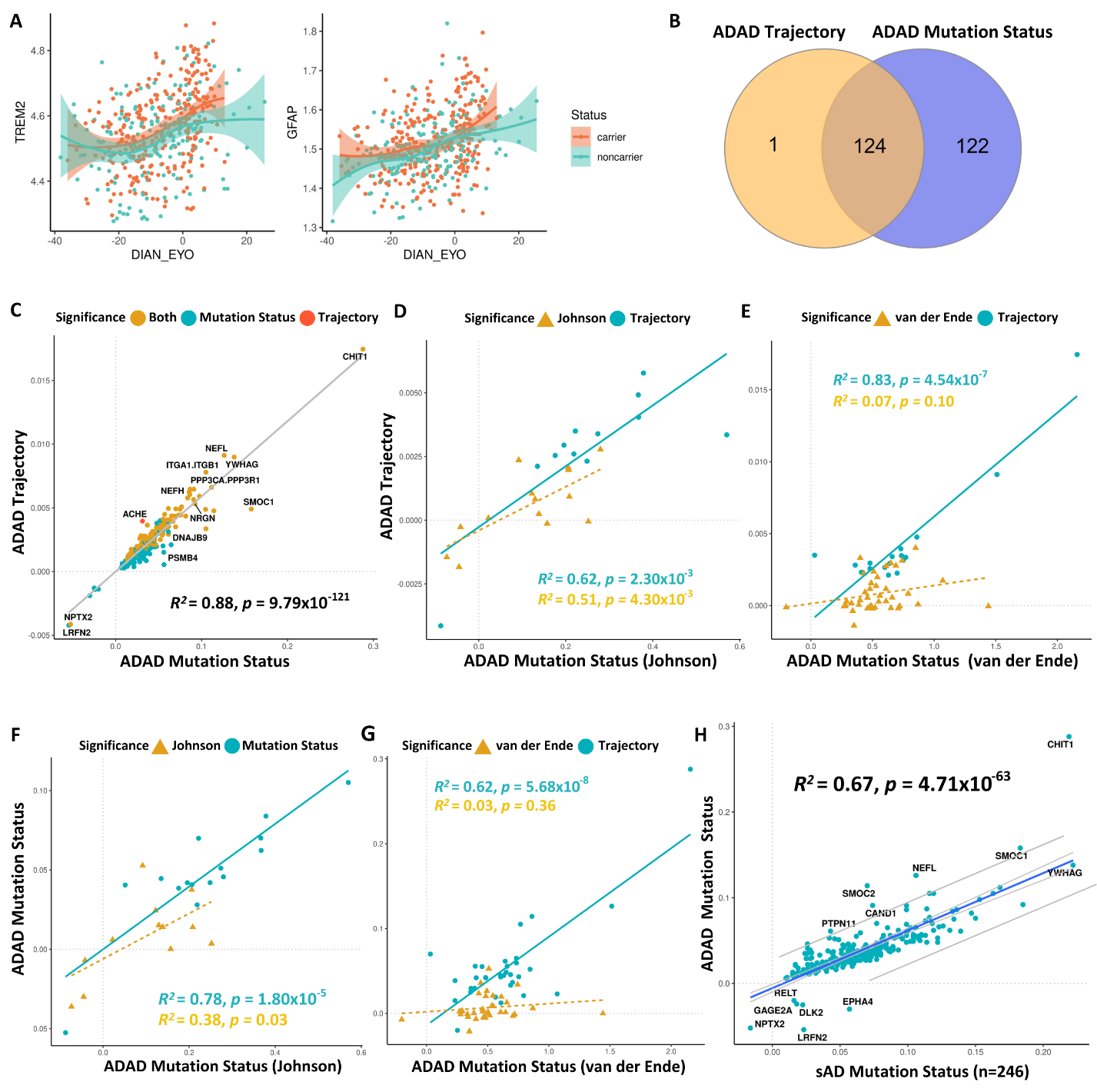
**Supplementary Figure 1**

**Supplementary Figure 1, The validation and replication for significant pseudo-trajectory proteins.** (A) The pseudo-trajectory curve for TREM2 and GFAP, but these two proteins did not pass the Bonferroni threshold between MCs vs.NCs. The red curve indicates the MCs proteins change with EYO, green curve indicates the NCs proteins change with EYO; (B) the overlapped significant proteins from trajectory analysis and ADAD mutation status analysis of MCs vs. NCs at Bonferroni threshold; (C-H) Scatterplot of the significant trajectory proteins replicated in (C) CSF ADAD mutation status analysis; (D) significant pseudo-trajectory proteins replicated in Johnson et al. finding; (E) significant trajectory proteins replicated in van der Ende et al. finding; (F) significant proteins associated with mutation status replicated in Johnson et al. finding; (G) significant proteins associated with mutation status replicated in van der Ende et al. finding; (H) significant proteins associated with mutation status replicated in significant proteins associated with sAD.

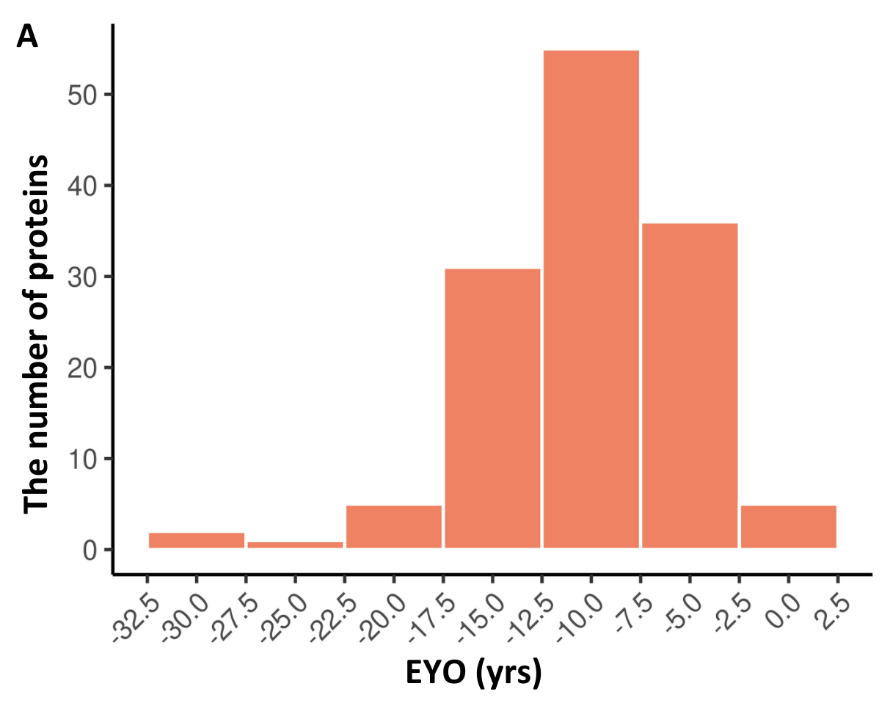
**Supplementary Figure 2**

**Supplementary Figure 2, Histograms of EYO distribution for 125** **significant trajectory proteins.** Each bar represents the number of significant proteins in the five years.

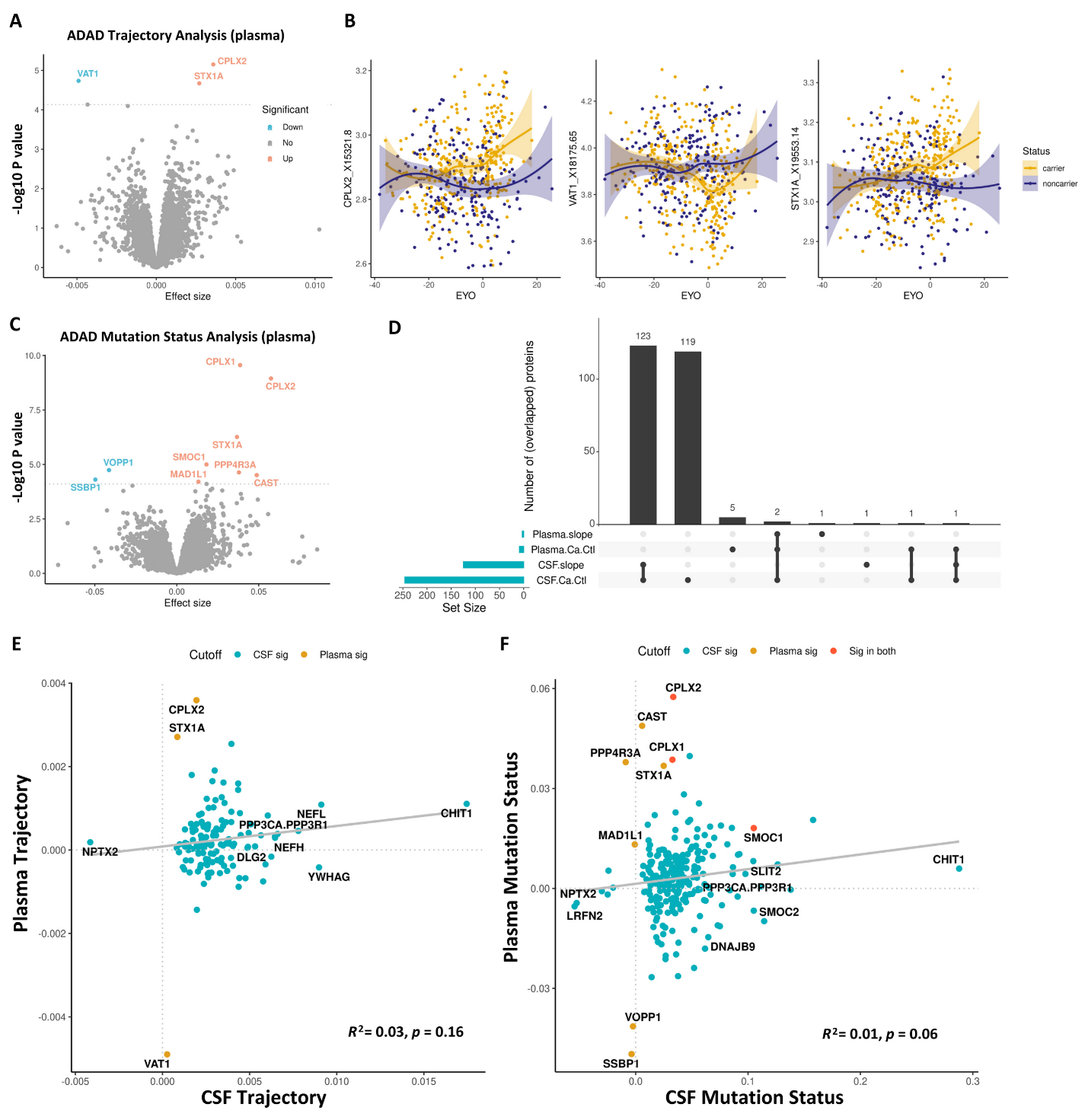
**Supplementary Figure 3**

**Supplementary Figure 3,** **The significant findings of trajectory and ADAD mutation status analysis from MCs vs. NCs in plasma.** (A-B) Volcano plots displaying the estimate change (x axis) against -log10 statistical differences (y axis) for all tested proteins. The red dots show the proteins upregulated significant proteins and the blue dots show the proteins downregulated significant proteins at FDR corrected threshold. (A) The significant trajectory proteins from MCs and NCs; (B) The significant proteins from ADAD mutation status analysis; (C) CPLX2 and STX1A showed the significant trajectory in plasma ; (D) Identify the overlapped significant proteins between CSF and plasma trajectory and ADAD mutation status analysis. The UpSet intersection diagram shows the overlapped proteins in the analysis; (E) The effect size correlations of significant trajectory proteins between CSF and plasma; (F) The effect size correlations of significant proteins between CSF and plasma ADAD mutation analysis.
**Supplementary Figure 4**

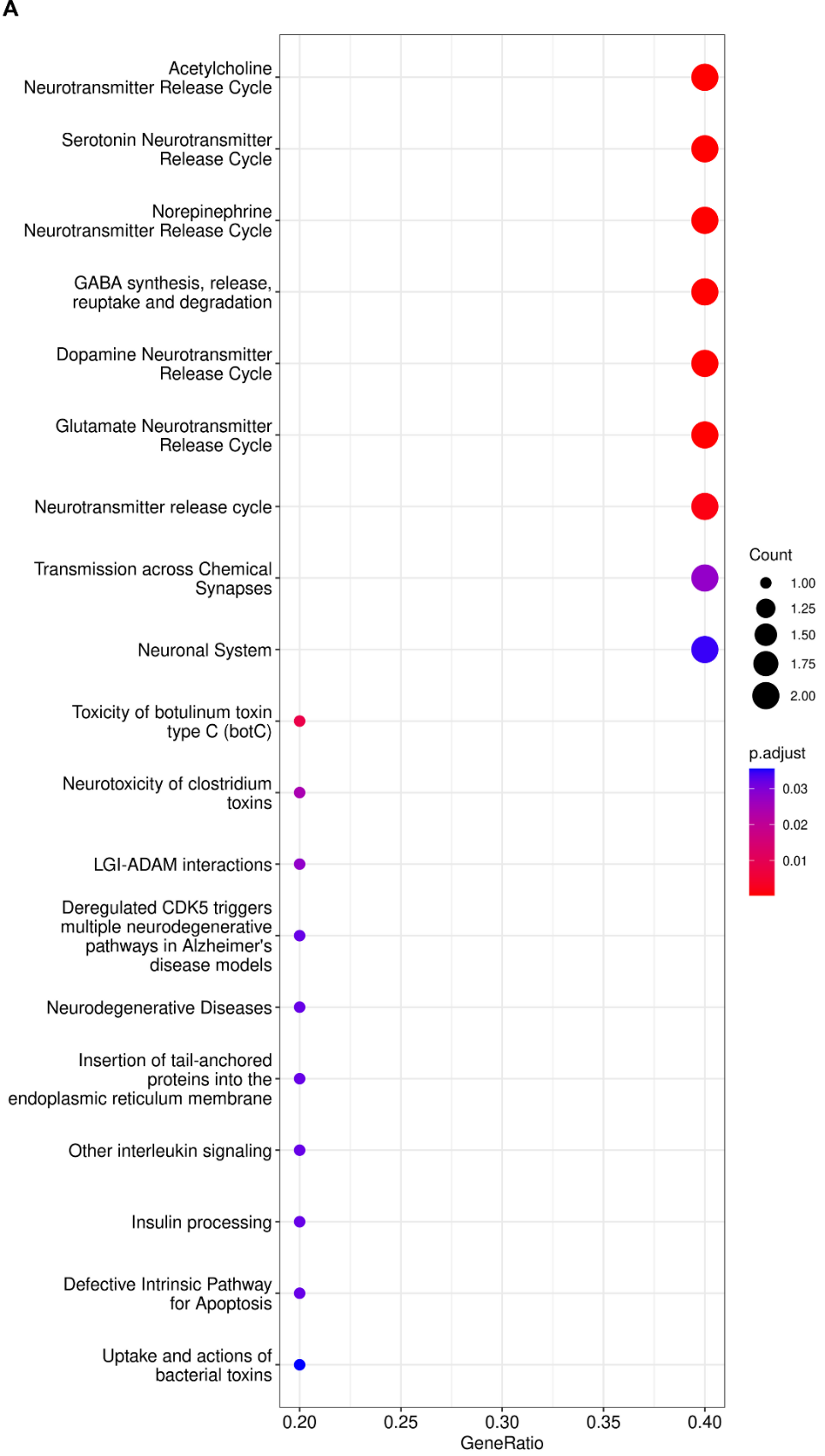

**Supplementary Figure 4, pathway enrichment analysis for significant proteins associated with ADAD status in plasma.** Enrichment listed the top 20 enriched pathways. The color from red to blue represents the FDR p value, from the lowest to the highest, respectively. The circle sizes indicate the gene count.

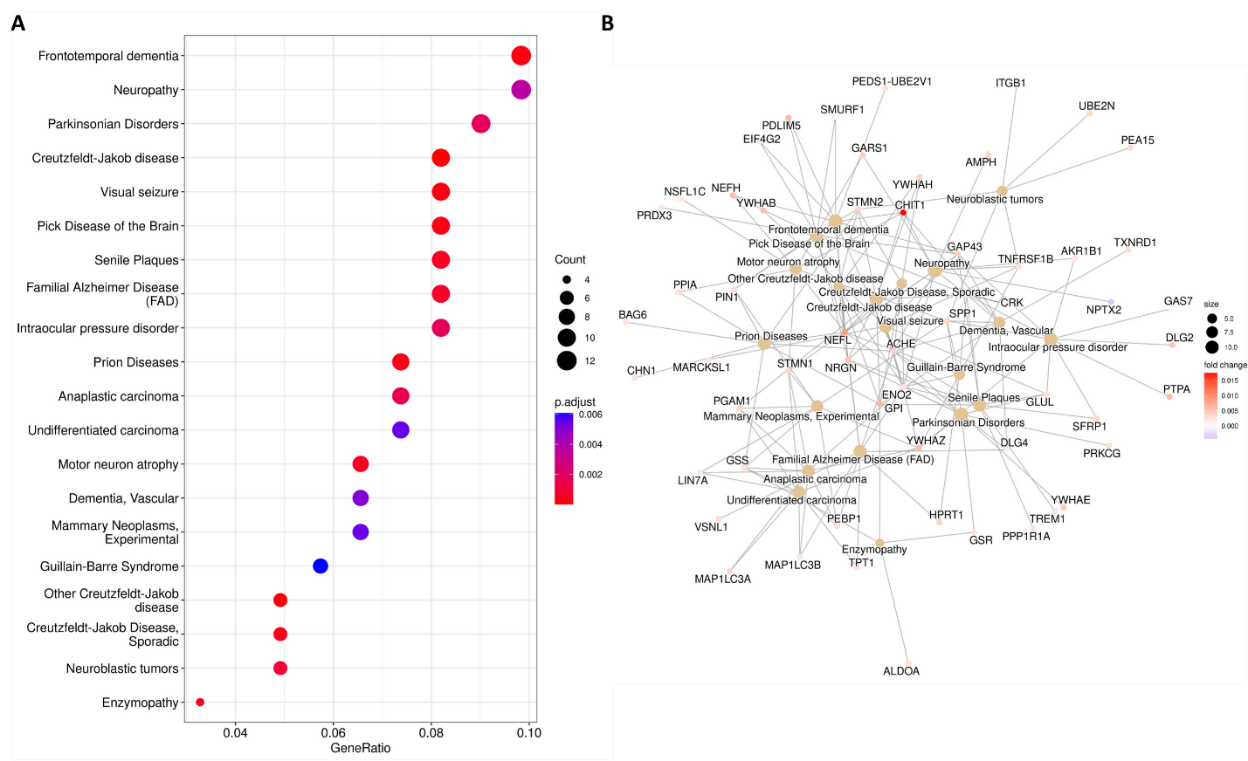
**Supplementary Figure 5**

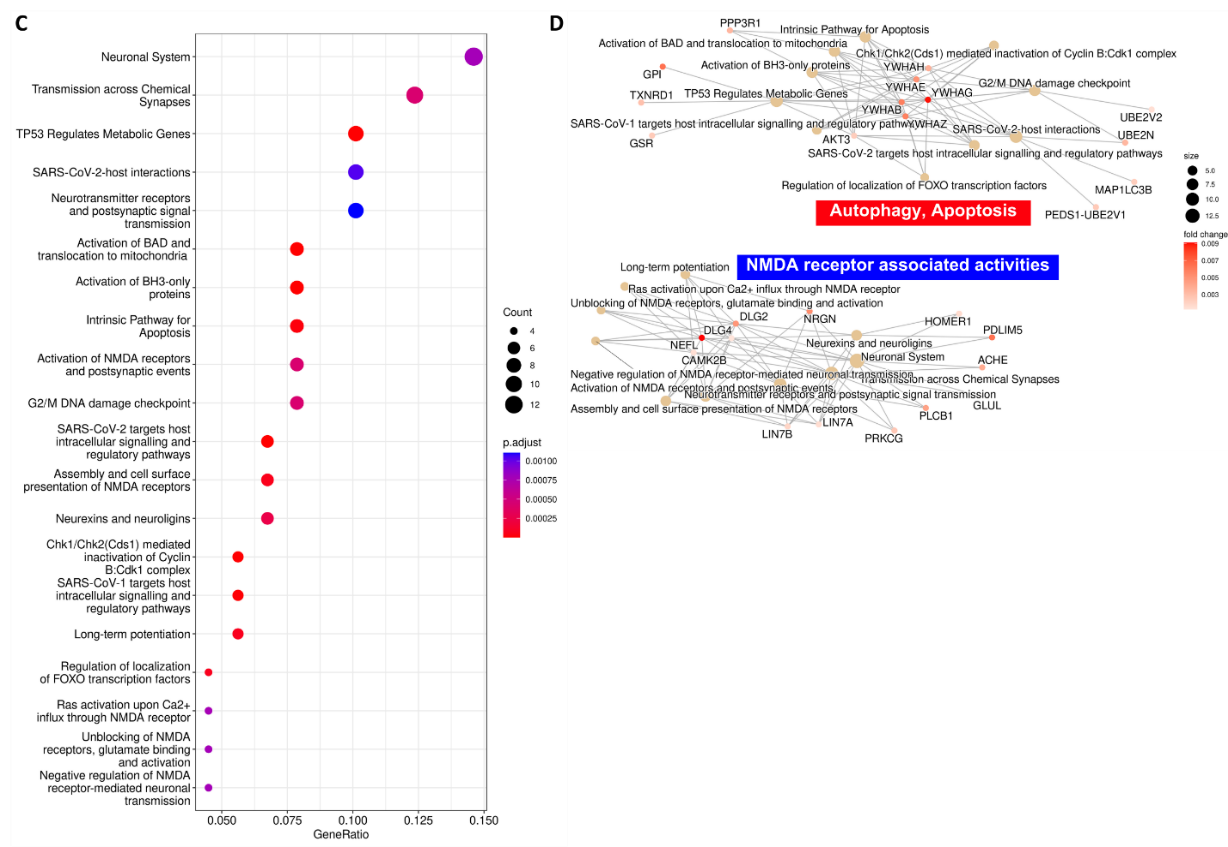

**Supplementary Figure 5, pathway enrichment analysis for significant trajectory proteins in CSF.** (A) Top 20 significant enriched diseases in DisGeNET enrichment analysis; (B) Enrichment map of the top 20 significant enriched diseases presented by the network;(C) Top 20 significant pathways in Reactome pathway analysis;(D) The top 20 significantly enriched Reactome pathways and their associated genes. Fold change was based on the estimate of each protein. The color from red to blue represents the FDR p value, from the lowest to the highest, respectively (A-C). The circle sizes indicate the gene count (A-D).

**Supplementary Figure 6**

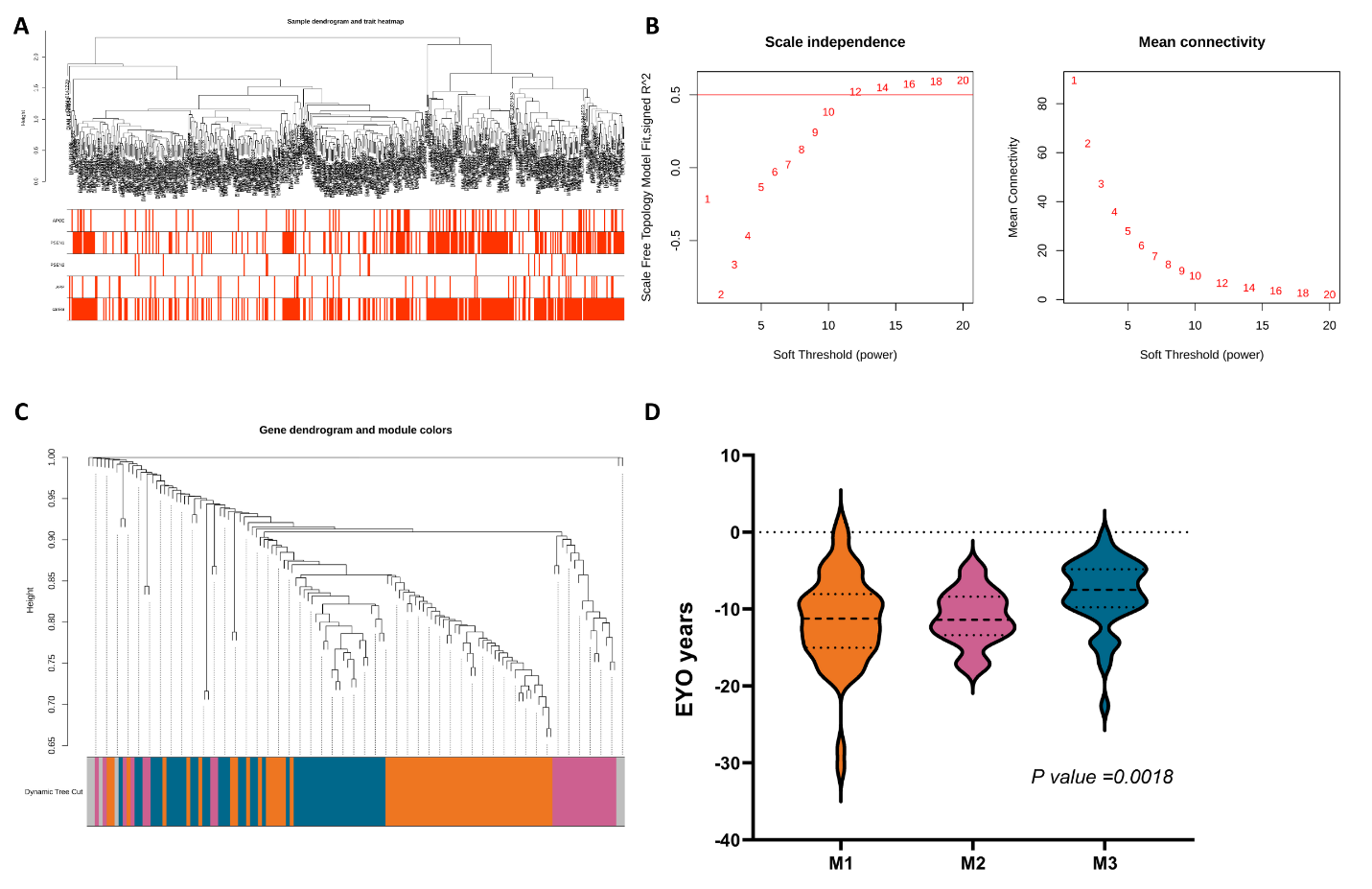

**Supplementary Figure 6, Construction of WGCNA.** (A) Sample dendrogram with traits heatmap. Red is ADAD in APOE E4, PSEN1,PSEN2,APP,Carriers. (B) The scale-free fit index and mean connectivity of WGCNA. It exhibits the soft thresholding power β in the WGCNA. The x-axis represents the soft-threshold power. Based on proportional independence and average connectivity analysis, β = 12 was selected as the soft threshold to construct the network. (C) Clustering dendrogram of proteins, dissimilarity is based on topological overlap, together with assigned module colors. As a result, 4 co-expression modules were constructed and was shown in different color. The number of proteins in each module were listed in Table S17.

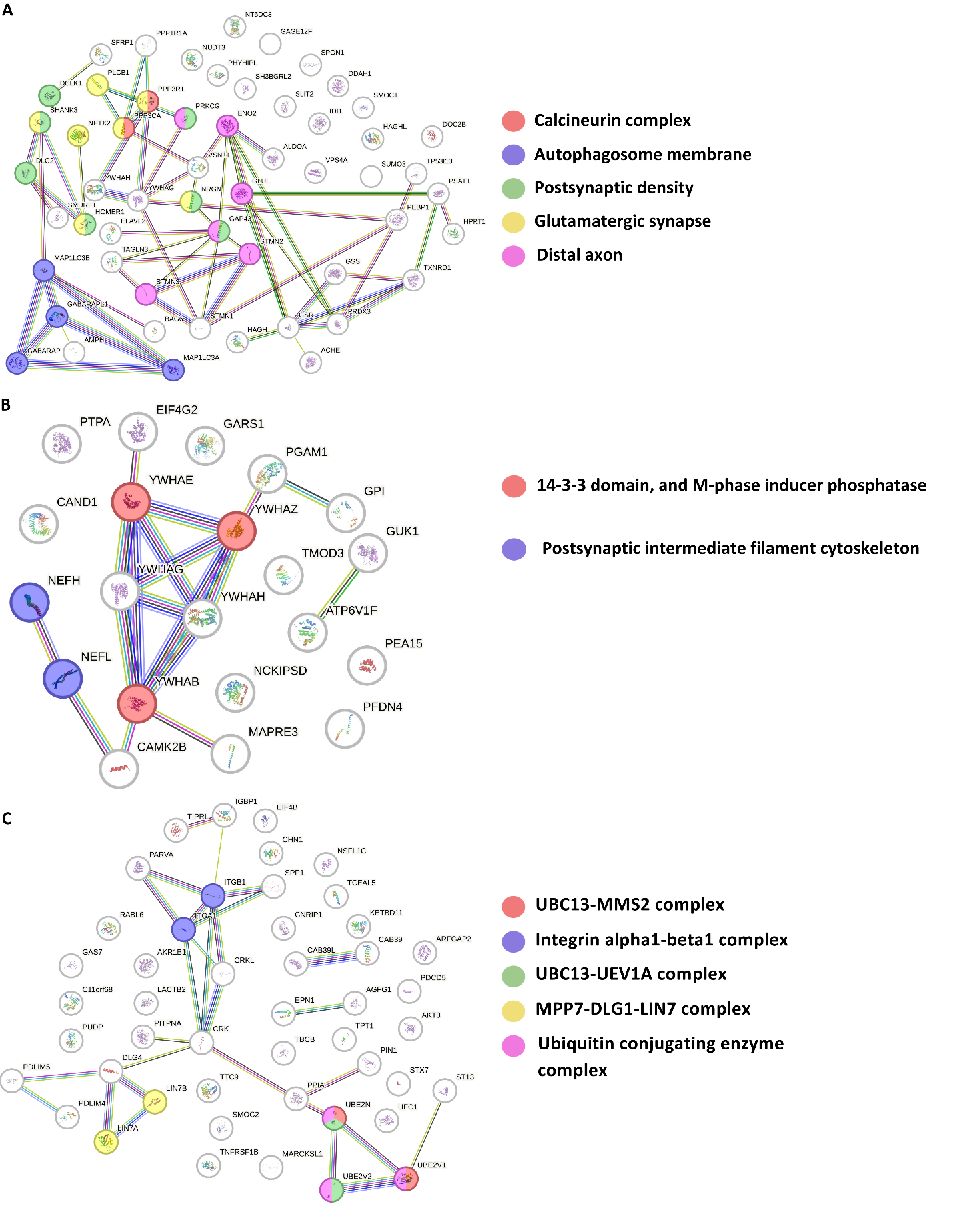
**Supplementary Figure 7**

**Supplementary Figure 7, PPI network analysis for each module.** (A) PPI network for M1 in GO cellular component. (B) PPI network for M2 in STRING network clusters. (C) PPI network for M3 in GO cellular component.
